# Diagnostic Feature Associations in Atypical Parkinsonian Disorders: A Survey of Movement Disorders Specialists

**DOI:** 10.64898/2026.11.02.26367134

**Authors:** Cole Zweber, Vivikta Iyer, Emily Houston, James T. Boyd, Lawrence Golbe, Michiko K. Bruno, Deepak K. Gupta

## Abstract

**Objective:** For the atypical parkinsonian disorders (APDs), including dementia with Lewy bodies (DLB), multiple system atrophy (MSA), progressive supranuclear palsy (PSP), and corticobasal degeneration (CBD), accurate antemortem diagnosis remains clinically challenging because of overlapping features, phenotypic heterogeneity, and limited definitive biomarkers. Consensus diagnostic criteria are complex and may not reflect how clinicians operationalize diagnostic reasoning in practice.

**Methods:** We surveyed movement disorders specialists regarding diagnostic implications of 32 clinical features, comparing the results with a 2024 consensus algorithmic framework called the “General Neurologist’s Practical Diagnostic Algorithm for APDs”. We evaluated inter-rater agreement, diagnosis-level framework coverage, feature-level attribution patterns, and exact-set agreement.

**Results:** Respondents demonstrated strong inter-specialist consensus in feature attribution (raw agreement 81.9%; Gwet’s AC1 0.67, 95% CI 0.64-0.70). Specialist endorsement increased markedly with framework association strength, from 12.9% to 36.2% to 82.6% for unmapped, modest, and moderate associations respectively. Diagnosis-level coverage was higher for MSA, PSP, and CBD than for DLB. Exact-set agreement was substantially more variable and depended on whether modest associations were incorporated into the framework definition. Despite high self-reported familiarity with and use of published diagnostic criteria, respondents overwhelmingly expressed interest in a unified diagnostic framework for APDs with automated clinical decision support (CDS).

**Conclusions:** Movement disorders specialists showed internally consistent feature-attribution patterns for archetypal features, while differing in the weighting and breadth assigned to weaker or overlapping associations. These findings are evidence for a gap between consensus schemas and specialist diagnostic reasoning and may inform future diagnostic frameworks and CDS approaches for the APDs.

## 1. INTRODUCTION

The atypical parkinsonian disorders (APDs), including dementia with Lewy bodies (DLB), multiple system atrophy (MSA), progressive supranuclear palsy (PSP), and corticobasal degeneration (CBD), share overlapping clinical signs and symptoms with one another and with Parkinson’s disease (PD), particularly early in the disease course ^1^. This overlap contributes to diagnostic uncertainty and misdiagnosis, which may delay appropriate counseling, management, and clinical trial enrollment. A 15-year longitudinal observational study of patients with Parkinsonian disorders identified frequent shifts in the leading diagnosis from PD to non-PD and the reverse, even multiple times in some cases ^2^.

Definitive diagnosis of parkinsonian disorders remains neuropathologic. Antemortem markers using fluids, tissue and imaging are highly promising but remain largely confined to research settings. As a result, diagnosis continues to rely on clinical consensus criteria for both PD ^3^ and APDs ^4–7^. These diagnostic criteria have categories such as “clinically probable” and “clinically established” with recent iterations incorporating a prodromal or “suggestive” phase ^4^ to promote early recognition.

However, this evolution exposes a fundamental tension: diagnostic sensitivity prioritized at early stages at the expense of specificity, while acceptable diagnostic certainty often emerges only after the optimal window for early intervention and trial participation has passed. In practice, clinicians must navigate this uncertainty using complex, multi-layered frameworks that are difficult to apply consistently at the point of care.

Efforts to improve feasibility of making tentative diagnoses for non-subspecialists include algorithmic approaches such as “A General Neurologist’s Practical Diagnostic Algorithm for Atypical Parkinsonian Disorders” (GNA) developed by the CurePSP Centers of Care^5^. While such tools acknowledge the need for diagnostic iteration and allow for overlapping or provisional diagnoses, they still present a complex manual task for the clinician at the point of care, limiting scalability, consistency, and real-time utility^6^.

While clinical disability scales for established disease such as the Movement Disorders Society Unified Parkinson’s Disease Rating Scale (MDS-UPDRS) have been implemented as simplified checklists in various electronic health records (EHR) platforms (e.g. Epic flowsheet), but diagnostic criteria present a more complex challenge, requiring capture of diverse clinical features and reliable application and interpretation of complex logical steps. This can be difficult to perform efficiently in routine care encounters or even in clinical trials, particularly when criteria are presented as text, tables, or multistep algorithms. Digital options for applying complex diagnostic criteria at the point of care in neurology remain highly limited. ^7^

Our prior work in PD (CDS-PD) has demonstrated the feasibility of CDS as support for applying the MDS-PD diagnostic criteria ^8^, but to our knowledge, there is currently no comparable CDS approach for APD criteria. Such an approach could facilitate simultaneous application of diagnostic frameworks for multiple APDs, an objective approximated by the GNA. However, the design of such tools requires an understanding of how expert clinicians currently associate and prioritize clinical features in practice.

To better understand this problem and to guide the future development of a digital CDS tool for the APDs, we conducted a survey of neurologists to understand how they associate individual history and examination features with DLB, MSA, PSP, and CBD. We also assessed respondents’ familiarity with and use of established APD diagnostic criteria and the published GNA. Our first steps were to characterize how movement disorders specialists associate clinical features with APD diagnoses and to explore similarities and differences between these diagnostic cognitive models and those recommended by a published diagnostic framework. We hypothesized that specialists would demonstrate recognizable but heterogeneous patterns of feature attribution and measured the degree to which these patterns overlapped with the GNA-derived mapping. We also tested the hypothesis that despite high claimed familiarity with individual APD diagnostic criteria, clinicians would report little formal reliance on these criteria in clinical practice.

## 2. MATERIALS AND METHODS

### 2.1 Survey Development and Distribution

We developed a survey using the Qualtrics platform hosted by the University of Vermont and, in a collaborative and iterative fashion among the coauthors, piloted it with two movement disorders neurologists at UVM Medical Center. The resulting version was approved by the UVM Institutional Review Board (IRB) under an exemption based on the absence of any plan to collect or distribute sensitive, identifiable information.

The survey was distributed to movement disorders neurologists, general neurologists, and other clinicians via the contact lists of four organizations: (1) CurePSP (through their Centers of Care email listserv); (2) American Academy of Neurology (through their online Synapse forum for Movement Disorders); (3) Parkinson Study Group (through their Atypical Parkinsonian Disorders working group’s email listserv); and (4) Michael J. Fox Foundation for Parkinson’s Research (through their Parkinson’s Disease Research Exchange platform). The survey was open from October 20, 2025, to March 1, 2026.

The survey comprised three sections: (1) Demographics; (2) Clinical Questionnaire; and (3) Familiarity and Clinical Use. The invitation, the informed consent information, and the survey itself appear as Supplementary Material.

### 2.2. Demographics

In this section, respondents reported their professional role (e.g., movement disorders neurologist, behavioral neurologist, other neurologist, non-neurologist physician, fellow, nurse practitioner, physician assistant), years in practice, and when applicable, years of fellowship training completed.

### 2.3 Clinical Questionnaire

This section required respondents to categorize the diagnostic utility of 32 clinical features (comprising 14 history-based and 18 examination-based features) derived from the GNA ^5^ (Supplementary Table 1). Specifically, for each feature, respondents selected any APD diagnoses (DLB, MSA, PSP, CBD) that were “supported by the presence of that feature,” assuming the feature was present early in the disease course, with multiple selections allowed. The corticobasal response option was presented to respondents as “corticobasal degeneration (CBD)” to align with the published consensus diagnostic criteria. However, as the GNA uses corticobasal syndrome (CBS), comparisons using the corticobasal category should be interpreted with caution and should not be taken to imply equivalence between the clinical syndrome and its underlying neuropathologic substrate. Additionally, respondents were not directed to consider a specific differential-diagnostic scenario (e.g. distinguishing APDs from PD, from one another, or from non-parkinsonian neurodegenerative disorders), nor were they asked to rank or weight the degree of diagnostic utility. Responses therefore reflect clinician-perceived associations between features and diagnoses rather than formal estimates of diagnostic sensitivity, specificity, or overall diagnostic performance. This parallels the approach taken by the GNA itself.

### 2.4 Familiarity and Clinical Use

The final section of the survey asked respondents to report whether they were familiar with the four individual APD diagnostic criteria and the GNA (No, Yes). For each set of diagnostic criteria/algorithm with which they were familiar, respondents were additionally prompted to report their frequency of use in patients in whom the relevant APD was suspected (0–5%, 6–50%, 51–95%, or >95% of the time). Lastly, respondents were asked to indicate their personal likelihood of using an EHR-based application to assist with APD criteria application if made available (Not likely, Likely, Very likely) and whether there is an overall need to develop a unified diagnostic schema combining separate, disease-specific sets of criteria into a probabilistic differential diagnosis (Unsure, No, Yes).

### 2.5 Data Analysis

De-identified Qualtrics survey data were exported as comma-separated values (CSV) and analyzed using Python in Google Colab ^9^. Figures were created using RStudio ^10^.

Analyses were conducted on completed surveys; incomplete surveys, defined as lacking entries for any of the mandatory questions in the Demographics and Familiarity and Clinical Use sections, were excluded from the analysis. Notably, non-selection of a diagnosis for a given clinical feature in the Clinical Questionnaire section did not constitute an incomplete response because respondents were permitted to indicate that none of the four APDs were supported by the feature. This was intended to allow respondents to express clinical discernment.

Primary analyses were restricted to respondents who identified as either “Movement Disorders Neurologist” or “Movement Disorders Fellow.” Secondary analyses were performed using response data from non-movement disorders specialist survey respondents, including “Behavioral neurologists,” “Other neurologists” and “Nurse practitioners.”

Descriptive statistics and frequency distributions were performed to characterize respondent demographics, feature-diagnosis attribution patterns, and familiarity and utilization of diagnostic criteria. For each clinical feature-diagnosis combination, we calculated the number and percentage of respondents selecting that diagnosis.

Inter-specialist agreement was evaluated independently of the published comparison framework. Each of the 128 possible feature-diagnosis combinations (32 clinical features × four APD diagnoses) was treated as a binary rating item evaluated by all 31 respondents. Raw percent agreement and Gwet’s AC1 with 95% confidence intervals were calculated for the complete set of feature-diagnosis pairs and separately for each diagnosis. Gwet’s AC1 was selected as a chance-corrected agreement coefficient suitable for binary ratings in which response prevalence may be highly imbalanced.

To compare respondent feature-attribution patterns with those represented in the GNA, we constructed a reference mapping from the feature-diagnosis associations specified in the published framework for each APD. All 128 clinical feature-diagnosis combinations were classified according to the strength assigned in the published framework: 0 for an association not represented in the framework, 1+ for a “modest predictive value” association, and 2+ for a “moderate predictive value” association (Supplementary Table 1). Notably, three of the 32 clinical features (“Major vascular risk factors or stroke history,” “Recent or current neuroleptic or other dopamine-blocking drug” and “Gait apraxia/frontal gait syndrome”) were not mapped as 2+ to any of the four APDs assessed in the survey, as they are instead most strongly associated with vascular parkinsonism, drug-induced parkinsonism, and normal pressure hydrocephalus, respectively. However, “Gait apraxia/frontal gait syndrome” was mapped as 1+ to both PSP and CBD. The resulting reference mapping contained 30 clinical features that were mapped as at least 1+ to one or more of the APDs, with 2 clinical features remaining unmapped.

Specialist endorsement was summarized at the individual feature-diagnosis level according to framework association strength. Diagnosis-level framework coverage was additionally calculated for each respondent as the proportion of features mapped to a particular diagnosis that the respondent associated with that diagnosis. This measure was calculated separately using moderate (2+) associations and all mapped (1+/2+) associations. Because coverage assessed recovery of framework-mapped associations, selection of additional diagnoses for an individual feature did not reduce coverage. Coverage distributions are reported as medians and interquartile ranges across the 31 specialist respondents.

Exact-set agreement was calculated as a more stringent secondary comparison. For each feature, a response was classified as an exact match only when the complete set of diagnoses selected by the respondent was identical to the diagnosis set defined by the framework. Both omission of a framework-mapped diagnosis and selection of an additional diagnosis resulted in classification as non-agreement. Exact-set agreement was calculated using two framework definitions: moderate associations only (2+ mapping), and all modest or moderate associations (1+/2+ mapping). When no diagnosis among the four APDs was mapped to a feature under the relevant definition, the reference set was empty and was matched only when the respondent selected no APD. Feature-level estimates are reported with 95% Wilson score confidence intervals. Respondent-level exact-set agreement was calculated as the percentage of the 32 clinical features for which each respondent reproduced the complete framework-defined diagnosis set, and distributions under the two mapping definitions were compared descriptively.

Because the survey presented corticobasal degeneration (CBD) as a diagnostic response option whereas the GNA uses corticobasal syndrome (CBS), the exact-set analyses were repeated after removing the corticobasal diagnostic category from both respondent selections and framework derived reference sets as a sensitivity analysis.

## 3. RESULTS

### 3.1 Demographics

Overall response rate could not be calculated due to overlap in organizational mailing lists and lack of access to unique recipient counts. We received 75 total survey responses, of which 45 (60.0%) were completed in full. Thirty-one completed responses were from movement disorders neurologists (n = 28) or fellows (n = 3) and constituted the primary analytic cohort. Mean experience was 12.06 ± 8.68 years, ranging from 1 year (a first-year movement disorders fellow) to 36 years.

### 3.2 Clinical Feature Attribution and Inter-Specialist Agreement

The top ten clinical features most frequently associated with each APD by movement disorders specialist respondents are shown in Table 1. For MSA, PSP, and CBD, the five most frequently selected features were each endorsed by more than 93% of respondents. In contrast, DLB showed less consistent feature attribution beyond the two most frequently selected features (“Dream enactment without recall (RBD)” and “Visual hallucinations not otherwise explained”) which were unanimously associated with DLB by all respondents.

**Table 1:**
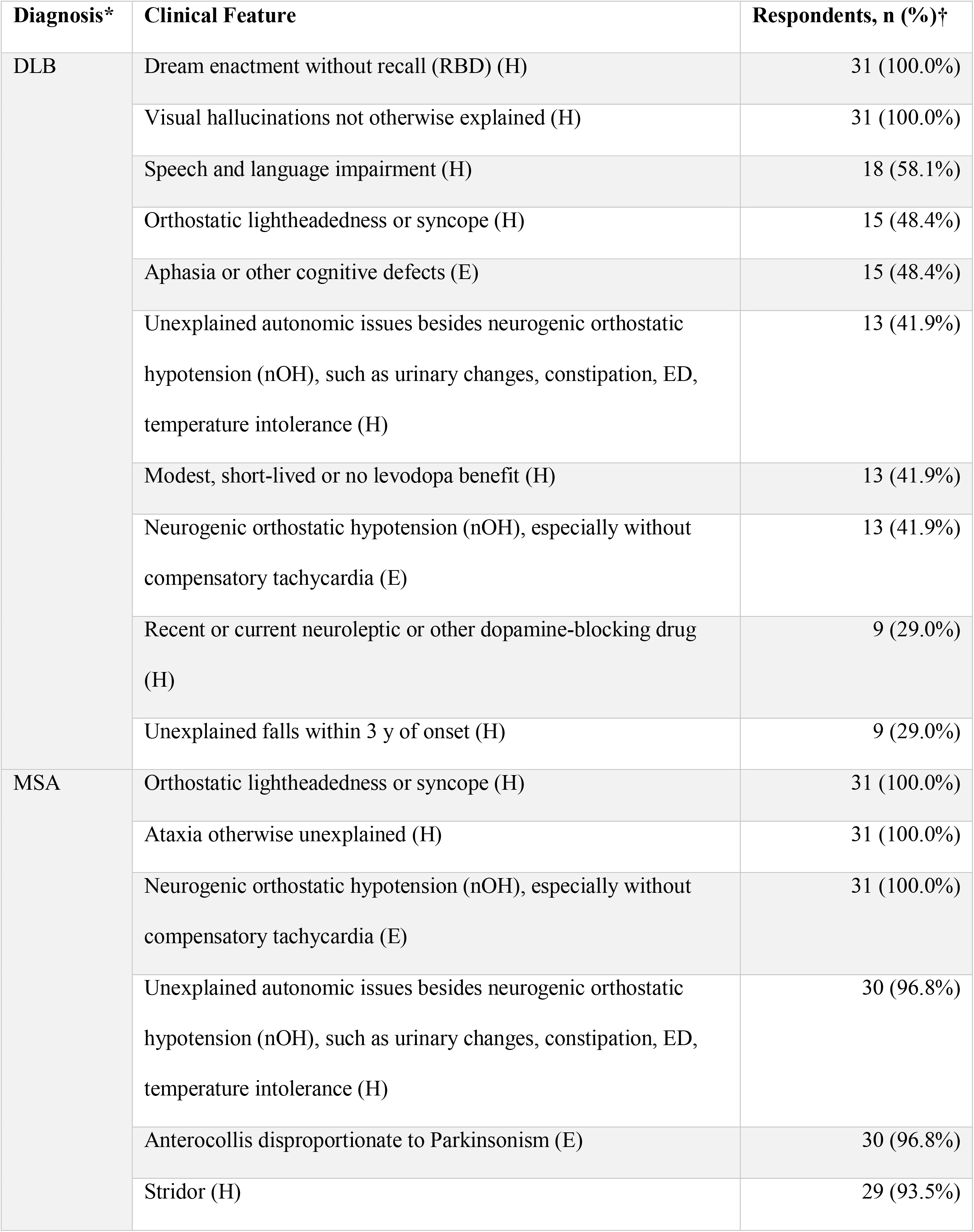
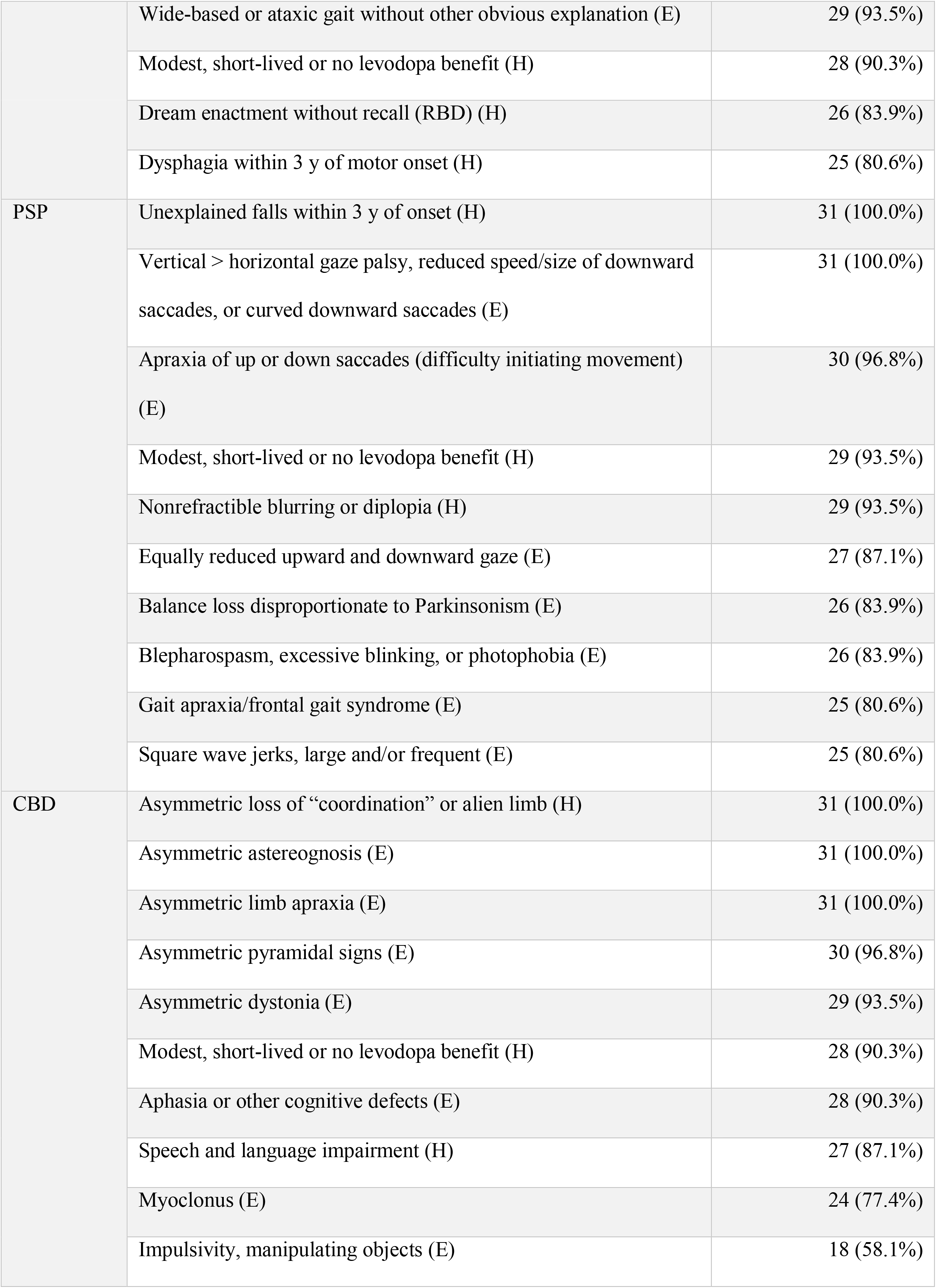
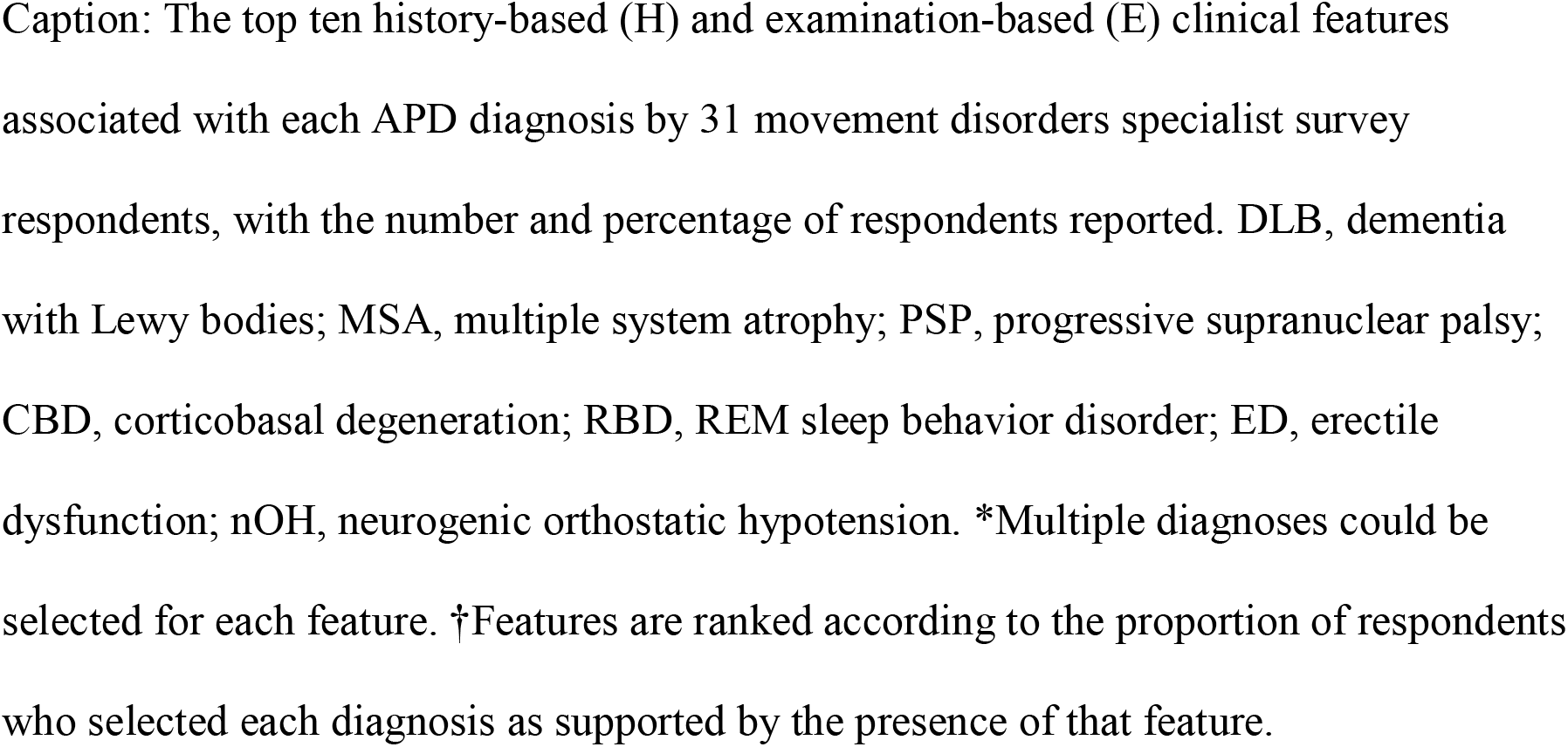
Clinical features most frequently associated with each APD diagnosis.

Independently of comparison with the published framework, overall raw agreement among specialists across the 128 feature-diagnosis judgements was 81.9%, with Gwet’s AC1 of 0.67 (95% CI, 0.64-0.70). Diagnosis-specific AC1 estimates were 0.75 (95% CI, 0.68-0.83) for DLB, 0.67 (0.61-0.73) for MSA, 0.62 (0.57-0.66) for PSP, and 0.64 (0.58-0.71) for CBD (Supplementary Table 2).

### 3.3 Relationship Between Specialist Attribution and Framework Association Strength

Across the 128 possible clinical feature-diagnosis combinations, 69 were not represented in the GNA for the corresponding diagnosis, 26 were designated as modest (1+) associations, and 33 were designated as moderate (2+) associations. Specialist endorsement increased markedly across these graded framework categories (Figure 1).

**Figure 1:**
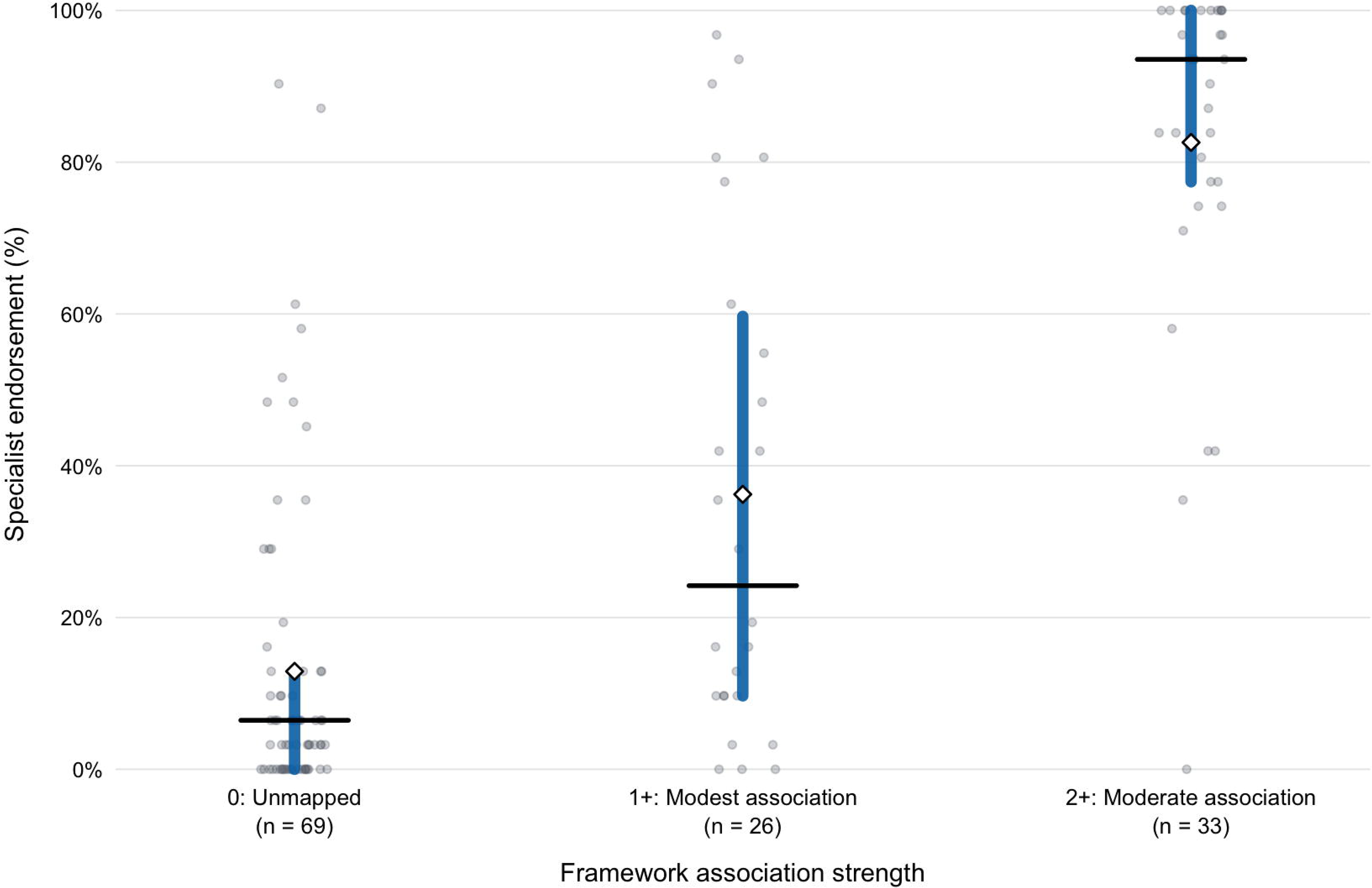
Specialist endorsement according to framework association strength. Caption: Each point represents one unique clinical feature-diagnosis pair among 128 possible combinations (32 clinical features × four APD diagnoses). Framework association strength was categorized as 0 (unmapped; n = 69), 1+ (modest association; n = 26), or 2+ (moderate association; n = 33). The vertical axis represents the percentage of 31 movement disorders specialist respondents who associated each feature with the corresponding diagnosis. Thick vertical lines indicate the interquartile range; horizontal black lines indicate the median; open diamonds indicate the mean.

Mean endorsement was 12.9% for unmapped associations, 36.2% for modest associations, and 82.6% for moderate associations; corresponding median endorsement values were 6.5%, 24.2%, and 93.5%, respectively. Individual estimates for all 128 feature-diagnosis combinations, with 95% confidence intervals, are provided in Supplementary Table 3.

Diagnosis-level analyses similarly showed greater coverage of moderately (2+) associated features than of the broader set of all mapped (1+/2+) associations (Table 2). Under the moderate (2+) mapping, median coverage was 50.0% (IQR, 50.0-66.7) for DLB and 88.9% for MSA, PSP, and CBD, with respective IQRs of 88.9-100.0, 77.8-100.0, and 77.8-100.0. When all modest and moderate (1+/2+) associations were incorporated, median coverage was 41.7% (29.2-50.0) for DLB, 63.2% (57.9-68.4) for MSA, 68.8% (62.5-81.2) for PSP, and 75.0% (66.7-75.0) for CBD.

**Table 2:** Diagnosis-level coverage of framework-mapped clinical features.

| **Diagnosis** | **Moderate (2+) associations, n** | **Coverage, median (IQR)** | **All mapped (1+/2+) associations, n** | **Coverage, median (IQR)** |
|---|---|---|---|---|
| **DLB** | 6 | 50.0% (50.0–66.7) | 12 | 41.7% (29.2–50.0) |
| **MSA** | 9 | 88.9% (88.9–100.0) | 19 | 63.2% (57.9–68.4) |
| **PSP** | 9 | 88.9% (77.8–100.0) | 16 | 68.8% (62.5–81.2) |
| **CBD** | 9 | 88.9% (77.8–100.0) | 12 | 75.0% (66.7–75.0) |

### 3.4 Exact-Set Agreement and Sensitivity Analyses

Exact-set agreement provided a substantially more stringent measure because respondents were required to reproduce all and only diagnoses represented in the framework-defined set for each feature. Feature level results under the moderate (2+) and all-mapped (1+/2+) definitions are provided in Supplementary Tables 4 and 5, respectively.

At the respondent level, median exact-set agreement was 56.3% (IQR, 45.3-59.4) using the moderate (2+) association definition, and 31.3% (IQR, 28.1-35.9) using the all-mapped (1+/2+) definition, with substantial inter-rater variation observed under both definitions (Figure 2).

**Figure 2:**
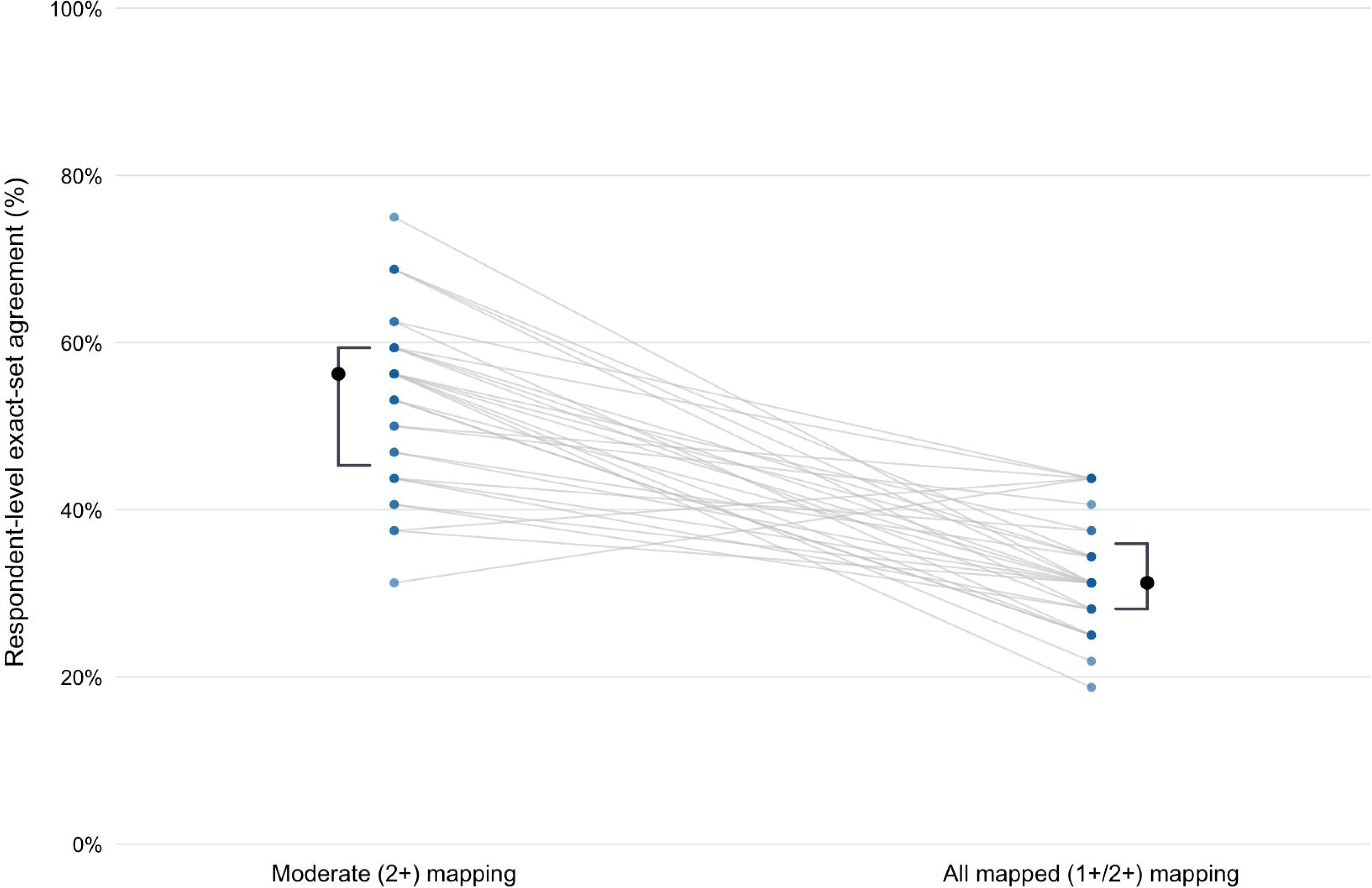
Distribution of respondent-level exact-set agreement under moderate and all-mapped framework definitions. Caption: Each paired set of points represents one of 31 movement disorders specialist respondents. For each respondent, exact-set agreement represents the percentage of 32 clinical features for which the set of APD diagnoses selected by the respondent was identical to the framework-defined diagnosis set. Both omission of a framework-mapped diagnosis and selection of an additional diagnosis resulted in non-agreement. Values are shown separately using moderate (2+) associations and all mapped (1+/2+) associations. Horizontal lines indicate the median and vertical brackets indicate the interquartile range. Thin connecting lines link estimates from the same respondent.

Feature-specific patterns also varied according to framework association. “Apraxia of up or down saccades (difficulty initiating movement),” the complete framework designated CBD as a moderate association and PSP as a modest association. Despite this, specialist respondents associated this feature substantially more often with PSP (96.8%) than with CBD (35.5%). “Equally reduced upward and downward gaze” was predominantly attributed to PSP (87.1%) despite PSP not being mapped to that feature as either moderate (2+) or even modest (1+) in the comparator framework (Supplementary Table 3).

Because the survey used CBD whereas the comparator framework uses CBS, exact-set analyses were repeated after removing the corticobasal diagnostic category from both respondent selections and reference sets. Feature-level sensitivity results are provided in Supplementary Table 6. Although individual feature estimates changed when the corticobasal dimension was removed, the broader finding that exact-set correspondence depended strongly on the mapping definition persisted.

### 3.5 Secondary Analysis of Non-Movement Disorders Specialist Respondents

The core analyses were repeated for the secondary cohort of respondents who did not identify as movement disorders specialists (n = 14). This secondary cohort was comprised of four “Behavioral neurologists”, eight “Other neurologists”, and two “Nurse practitioners”. Independently of comparison with the published framework, overall raw agreement among non-movement disorders specialist respondents was 71.5% with Gwet’s AC1 of 0.53 (95% CI, 0.49-0.56). Diagnoses specific AC1 estimates were 0.45 (95% CI, 0.40-0.50) for DLB, 0.62 (0.54-0.70) for MSA, 0.47 (0.41-0.53) for PSP, and 0.55 (0.48-0.62) for CBD (Supplementary table 7).

As with the main analytic cohort, diagnosis-level analyses showed greater coverage of moderately (2+) associated features than of the broader set of all mapped (1+/2+) associations in the non-movement disorders specialist respondents (Supplementary table 8). At the respondent level, among non-movement disorders specialist respondents, median exact-set agreement was 37.5% (IQR, 25.0-53.1) using the moderate (2+) association definition, and 17.2% (IQR, 10.2-31.3) using the all-mapped (1+/2+) definition.

### 3.6 Familiarity, Utilization, and Attitudes Toward Clinical Decision Support

Supplementary Figure 1 shows familiarity with the individual APD diagnostic criteria and the GNA, and Supplementary Figure 2 shows self-reported utilization. Awareness of the four individual sets of APD criteria was high (≥87% for each) with MDS-MSA criteria being highest (96.8%), followed by the DLB consortium criteria and the MDS-PSP criteria (each 93.5%), and then the CBD criteria (87.1%). For each of the four individual APD criteria, self-reported clinical usage exceeded 58.1% of respondents, the greatest being the MDS-MSA criteria at 80.6%. Awareness of the GNA was lower (58.1%), and even fewer (29.0%) reported using the GNA in >50% of cases in which an APD was suspected.

Reported personal likelihood of using an EHR-based application is shown in Supplementary Figure 3, and opinions regarding the need for a unified diagnostic schema are shown in Supplementary Figure 4. Among the 31 movement disorder specialist respondents, more than 80% reported that they would be inclined to use an EHR-based application to assist with criteria application (32.3% “Very likely”. 48.4% “Likely”). In addition, 77.4% of respondents indicated that a unified diagnostic schema is needed to assist APD diagnosis, with an additional 16.1% “Unsure”.

## 4. DISCUSSION

### 4.1 Key Findings and Diagnosis-Specific Patterns

In this survey of movement disorders specialists, we identified three complementary features of diagnostic attribution. First, specialists demonstrated appreciable inter-respondent consistency in assigning individual clinical features to APD diagnoses independently of any external framework. Second, specialist endorsement showed a strong graded relationship with association strength in the GNA with moderate (2+) associations endorsed far more frequently than modest (1+) or unmapped associations. Third, exact reproduction of complete-framework defined diagnosis sets was substantially more variable and sensitive to whether modest (1+) associations were incorporated. Together, these findings suggest that specialists and the comparator framework share a recognizable diagnostic core while differing in the breadth and weighting assigned to overlapping or less strongly represented associations. The secondary cohort analysis provides additional context for this interpretation. Movement disorders specialists demonstrated greater overall inter-rater agreement than non-specialist respondents (81.9% vs 71.5% raw agreement), consistent with the idea that subspecialty training contributes to more consistent feature-attribution in APDs. While this secondary cohort was smaller and more heterogeneous, the finding supports the interpretation that the observed attribution patterns in this study do not reflect random variation in feature selection.

Diagnosis-specific attribution patterns provide additional insight into the clinical domains in which this shared specialist framework is most apparent, as well as those in which greater variability emerges. Visual hallucinations and RBD were unanimously associated with DLB by movement disorders specialists, consistent with their prominence in established diagnostic criteria ^11^. Although fewer additional features were frequently attributed to DLB than to MSA, PSP, or CBD, this pattern did not reflect low overall inter-specialist agreement; diagnosis-specific agreement was numerically highest for DLB (AC1 0.75). Rather, DLB showed comparatively lower correspondence with both feature sets derived from the comparator framework. This distinction suggests a relatively consistent mental model centered on core DLB manifestations, while the broader organization of DLB-associated features differed from that represented in the comparator framework. This discrepancy may be due to the challenges differentiating between DLB and PD dementia (PD-D) as overlapping neuronal α-synucleinopathies (as opposed to the glial α-synucleinopathy of MSA), a diagnostic consideration exacerbated by the omission of PD and PD-D from the survey response options. Alternatively, it may reflect DLB’s cross-disciplinary nature, which is commonly managed by both movement disorder specialists and behavioral neurologists. Notably, the secondary cohort of non-movement disorders specialists demonstrated higher median coverage of framework-mapped DLB features at the diagnosis-level under both the moderate (58.3% compared to 50.05%) and all-mapped (45.8% compared to 41.7%) thresholds, lending some credence to a cross-disciplinary model of expertise.

For MSA, respondents demonstrated high consensus for core features of the MDS-MSA criteria ^12^ such as autonomic dysfunction and cerebellar syndrome. Notably, orthostatic hypotension was more consistently selected than other manifestations of autonomic dysfunction, such as urinary symptoms, consistent with greater salience of features perceived as more specific, measurable, or traditionally emphasized in diagnostic frameworks.

For PSP, respondents most consistently chose features characteristic of Richardson syndrome (PSP-RS)—ocular motor dysfunction and postural instability—core features of PSP-RS in the MDS-PSP criteria ^13^. In contrast, features associated with alternative PSP subtypes (e.g., predominant parkinsonism or frontal cognitive dysfunction) were selected less consistently by respondents. Among clinically supportive features, levodopa resistance (CC1) was frequently endorsed, whereas dysphagia (CC3) was less consistently selected. These findings suggest preferential weighting of archetypal disease representations during diagnostic reasoning, whether through greater familiarity with PSP-RS phenotype, or perception of its features as more diagnostically discriminative.

For CBD, asymmetric motor and cortical features were prominent, with the top five most frequently associated features reflecting lateralized motor or higher-order cortical dysfunction. Language and cognitive features were also frequently associated with CBD, including aphasia or other cognitive defects and speech and language impairment. Together, these response patterns are consistent with the salience of the classic asymmetric motor-cortical phenotype, while also reflecting the broader cognitive and language spectrum of corticobasal presentation ^14^.

### 4.2 Interpreting Framework Correspondence

Our use of the predefined feature-mapping of the GNA was intended to provide a single, clinically recognizable comparison framework rather than a validated diagnostic reference standard. The algorithm framework and clinician feature-attribution patterns observed in our survey can therefore be viewed as complementary representations of diagnostic knowledge. Examining similarities and differences between these approaches may provide insight into how structured frameworks correspond to specialist diagnostic reasoning in practice.

Feature-level exact-set agreement requires simultaneous reproduction of every diagnosis represented at a specified framework threshold while additionally penalizing diagnoses selected outside that set. Consequently, a specialist may identify a framework-mapped association yet fail to achieve an exact match because an additional clinically plausible diagnosis was also selected. The effect of association threshold on exact-set agreement was feature dependent. Inclusion of modest (1+) framework associations brought the framework-defined set closer to specialist attribution for some features, but required additional, inconsistently endorsed associations for others.

Diagnosis-level coverage and individual feature-diagnosis analysis address complementary questions by preserving recognition of mapped associations without automatically treating broader attribution as disagreement. Viewed together, these measures distinguish recognition of framework-supported associations from exact reproduction of the framework’s boundaries.

Specific discrepant associations further illustrate that multiple mechanisms underlie low exact-set agreement. “Apraxia of up or down saccades” was predominantly associated with PSP by respondents even though the strongest framework association was with the corticobasal category. Importantly, for this clinical feature, PSP was represented as a modest (1+) association in the complete framework. Conversely, “Modest, short-lived or no levodopa benefit” was broadly attributed across several APDs by respondents, while “Equally reduced upward and downward gaze” was predominantly attributed to PSP despite a substantially different pattern in the comparator framework. Together, these examples suggest that incomplete exact-set agreement can reflect both differences in framework thresholds and genuine differences in the relative diagnostic weight assigned to overlapping features.

### 4.3 Clinical Context of Feature Attribution

The combination of substantial inter-specialist agreement and graded correspondence with framework association strength suggests that specialist feature attribution is structured rather than idiosyncratic, albeit with incomplete overlap. Several factors may contribute to the observed differences between specialist feature attribution and the comparator framework and likely reflect the interplay of both cognitive and structural factors. Diagnostic reasoning commonly involves selective weighting of features, and while consensus criteria provide structured frameworks, clinicians do not uniformly apply them as rigid algorithms at the point of care. Instead, real-world diagnostic reasoning is often guided by heuristic, experience-based processes that generate internal mental models of disease shaped by training and experience, often emphasizing certain features or archetypal presentations. These tendencies were illustrated by the PSP findings, where respondents disproportionately favored features associated with PSP-RS. The strong attribution of aphasic and speech-language features to CBD also reflects the broader phenotypic spectrum relevant to corticobasal presentations. Progressive apraxia of speech and nonfluent/agrammatic language syndromes may precede the emergence of an overt parkinsonian phenotype and have been associated with tauopathies including CBS/CBD and PSP. Contemporary reports similarly describe primary progressive apraxia of speech evolving toward CBS- or PSP-like parkinsonian syndromes.^15, 16^ These syndromes illustrate how diagnostic representations may extend beyond the classic motor phenotype.

Additionally, formal diagnostic criteria and practical algorithms serve a different purpose from clinician mental representations: they are constructed through expert consensus processes that evolve over time and must standardize diagnostic evidence across users and settings and attempt to balance clinical usability with fidelity to underlying clinicopathologic relationships. Collectively, these interacting cognitive and structural influences may help explain why clinicians may demonstrate incomplete alignment with structured diagnostic schemas despite high familiarity, and with the most prominent discrepancies among overlapping or lower-weighted features rather than for archetypal findings.

### 4.4 Clinical Implications and Decision Support

Respondents reported high familiarity with and frequent usage of individual diagnostic criteria, but comparatively lower utilization of the integrated GNA. Consistent, real-time implementation of complex diagnostic frameworks and disease scales has been shown to be a challenge in clinical settings, with demonstrably poor retest reliability ^17, 18^.

Additionally, mixed pathologies and overlapping presentations are common in neurodegenerative disease ^19–22^, such that patients may meet criteria for multiple diagnoses, each with differing probabilities, a scenario not well addressed by current siloed diagnostic frameworks. While the GNA represents an attempt to apply a more unified approach, adoption among surveyed movement disorder specialists remains limited.

At the same time, most respondents indicated willingness to use an EHR-based application for criteria implementation and perceived a need for a unified APD diagnostic schema. The greater variability in weaker or overlapping associations, and demonstrable clinician interest in structured digital support tools provides a practical rationale for development of CDS capable of integrating multiple diagnostic frameworks while preserving clinician judgment.

### 4.5 Limitations and Future Directions

Several limitations should be considered. First, the primary analytic sample comprised 31 movement disorders specialists recruited through voluntary participation, introducing potential selection bias and limiting generalizability. Secondly, the survey measured feature-diagnosis attribution rather than diagnostic performance. Respondents were not given complete patient vignettes nor asked to consider a specific differential diagnostic context or the relative diagnostic strength of individual associations. As such, responses reflect clinicians’ mental associations between features and diagnoses rather than formal assessments of diagnostic sensitivity, specificity, or positive/negative predictive values. Third, the 32 clinical features selected from the GNA enabled parallel assessment of the four APDs but did not comprehensively reproduce the respective complete consensus criteria for each individual APD, or subtype specific features. This approach may introduce both potential loss of nuance and represents an algorithmic approach for which the intended audience was not movement disorders specialists, but rather general neurologists. Fourth, the survey used CBD whereas the GNA uses CBS. These constructs are not interchangeable: CBS is a clinical phenotype that may arise from CBD or other underlying pathologies, whereas CBD denotes a neuropathological entity with multiple possible clinical pictures. We attempted to mitigate this by performing sensitivity analyses; however, this discrepancy in terminology may have influenced feature attribution. Finally, the primary analytic cohort was restricted to movement disorders specialists. While our principal objective was to characterize subspecialist reasoning, it may have preferentially emphasized motor and movement-disorder constructs relative to cognitive or behavioral perspectives. This limitation is particularly relevant in DLB and CBS/CBD, in which behavioral neurologists may encounter and weight cognitive, language, and neuropsychiatric manifestation differently.

Future studies should also examine how specialist feature attribution changes when biological information is incorporated. Emerging α-synuclein, amyloid-β, phosphorylated tau, and other biomarkers increasingly complement phenotype-based classification of neurodegenerative disease, including frequent co-pathology with Alzheimer’s disease.

Accordingly, there is momentum towards reframing the classification and staging of parkinsonian disorders to include, and even hinge on the presence of biomarkers. ^23, 24^ Integrating these measures with structured clinical features may ultimately provide a more complete basis for diagnostic frameworks and future CDS than either clinical or biological information alone.

### 4.6 Conclusion

In summary, movement disorders specialists demonstrated internally consistent patterns of APD feature attribution that broadly tracked the graded association strengths represented in a published practical diagnostic framework. Correspondence was strongest for moderately associated and archetypal features, whereas weaker and overlapping associations showed greater variability in specialist attribution. These findings suggest that specialists share a recognizable diagnostic core while differing in how less characteristic features are weighted and integrated. Understanding these patterns may help inform future diagnostic frameworks and the prospective evaluation of CDS approaches for APD diagnosis.

## Data Availability

The dataset used in this publication will be made available by request from any qualified investigator. To make such a request, please contact the corresponding author of this paper from your institutional email address.

## ACKNOWLEDGEMENTS

We thank CurePSP, American Academy of Neurology, Parkinson Study Group, and Michael J. Fox Foundation for Parkinson’s Research for their collaboration and support in disseminating this study survey.

This study was funded by the project titled “Ontology-based, Real-time, Machine learning Informatics System for Parkinson’s Disease (ORMIS-PD)”, funded by the Early Investigator Research Award (EIRA) of the Parkinson Research Program (PRP) of the Congressionally Directed Medical Research Program (CDMRP) of the United States (US) Department of Defense (DOD). Award # W81XWH2110859.

