## Supplementary Materials for "Diagnostic Feature Associations in Atypical Parkinsonian Disorders: A Survey of Movement Disorders Specialists"

**SUPPLEMENTARY TABLES**

Supplementary Table 1: Clinical features included in the survey, as derived from the General Neurologist’s Algorithm with the associated strength of association for each diagnosis

| Clinical Feature | DLB | MSA | PSP | CBD |
| --- | --- | --- | --- | --- |
| Dysphagia within 3 y of motor onset (H) | 0 | 1+ | 2+ | 0 |
| Nonrefractible blurring or diplopia (H) | 0 | 1+ | 2+ | 0 |
| Unexplained falls within 3 y of onset (H) | 0 | 1+ | 2+ | 0 |
| Modest, short-lived or no levodopa benefit (H) | 2+ | 1+ | 1+ | 0 |
| Speech and language impairment (H) | 2+ | 0 | 1+ | 2+ |
| Stridor (H) | 0 | 2+ | 0 | 0 |
| Asymmetric loss of “coordination” or alien limb (H) | 0 | 0 | 0 | 2+ |
| Ataxia otherwise unexplained (H) | 0 | 2+ | 0 | 0 |
| Dream enactment without recall (RBD) (H) | 2+ | 2+ | 0 | 0 |
| Major vascular risk factors or stroke history (H) | 0 | 0 | 0 | 0 |
| Orthostatic lightheadedness or syncope (H) | 1+ | 2+ | 0 | 0 |
| Recent or current neuroleptic or other dopamine-blocking drug (H) | 0 | 0 | 0 | 0 |
| Unexplained autonomic issues besides neurogenic orthostatic hypotension (nOH), such as urinary changes, constipation, ED, temperature intolerance (H) | 1+ | 2+ | 0 | 0 |
| Visual hallucinations not otherwise explained (H) | 2+ | 0 | 0 | 0 |
| Balance loss disproportionate to Parkinsonism (E) | 1+ | 2+ | 2+ | 0 |
| Blepharospasm, excessive blinking, or photophobia (E) | 0 | 1+ | 2+ | 0 |
| Impulsivity, manipulating objects (E) | 0 | 1+ | 2+ | 0 |
| Equally reduced upward and downward gaze (E) | 2+ | 1+ | 0 | 1+ |
| Palilalia (E) | 0 | 0 | 2+ | 0 |
| Square wave jerks, large and/or frequent (E) | 0 | 1+ | 2+ | 0 |
| Vertical > horizontal gaze palsy, reduced speed/size of downward saccades, or curved downward saccades (E) | 0 | 0 | 2+ | 1+ |
| Gait apraxia/frontal gait syndrome (E) | 0 | 0 | 1+ | 1+ |
| Wide-based or ataxic gait without other obvious explanation (E) | 0 | 2+ | 1+ | 0 |
| Aphasia or other cognitive defects (E) | 0 | 0 | 0 | 2+ |
| Anterocollis disproportionate to Parkinsonism (E) | 1+ | 2+ | 0 | 0 |
| Apraxia of up or down saccades (difficulty initiating movement) (E) | 0 | 0 | 1+ | 2+ |
| Asymmetric dystonia (E) | 0 | 1+ | 1+ | 2+ |
| Asymmetric limb apraxia (E) | 0 | 0 | 1+ | 2+ |
| Asymmetric pyramidal signs (E) | 0 | 0 | 0 | 2+ |
| Myoclonus (E) | 1+ | 1+ | 0 | 2+ |
| Asymmetric astereognosis (E) | 1+ | 0 | 0 | 2+ |
| Neurogenic orthostatic hypotension (nOH) especially without compensatory tachycardia (E) | 2+ | 2+ | 0 | 0 |

Caption: The 32 clinical features included in the Clinical Questionnaire section of the survey. Features were derived from the General Neurologist’s Algorithm and classified according to source (history or examination). For each feature, the corresponding strength of association with each of the four APD diagnoses as per the General Neurologist’s Algorithm is provided, with 0 corresponding to no association, 1+ corresponding to “modest” association, and 2+ corresponding to “moderate” association. DLB, dementia with Lewy bodies; MSA, multiple system atrophy; PSP, progressive supranuclear palsy; CBD, corticobasal degeneration; RBD, REM sleep behavior disorder; ED, erectile dysfunction; nOH, neurogenic orthostatic hypotension.

Supplementary Table 2: Inter-rater agreement in feature-diagnosis attribution, overall and by diagnosis

| Analysis set | Feature-diagnosis pairs, n | Raw agreement (%) | Gwet's AC1 [95% CI] |
| --- | --- | --- | --- |
| Overall | 128 | 81.9 | 0.67 [0.64, 0.70] |
| DLB | 32 | 83.0 | 0.75 [0.68, 0.83] |
| MSA | 32 | 82.9 | 0.67 [0.61, 0.73] |
| PSP | 32 | 80.6 | 0.62 [0.57, 0.66] |
| CBD | 32 | 81.1 | 0.64 [0.58, 0.71] |

Caption: Inter-specialist agreement in binary feature-diagnosis attribution among 31 movement disorders specialist respondents. Each of the 128 feature-diagnosis pairs (32 clinical features × four APD diagnoses) was treated as an item rated independently by all respondents. Raw percent agreement and Gwet's AC1 with 95% confidence intervals are reported for the complete set of feature-diagnosis pairs and separately for each diagnosis. DLB, dementia with Lewy bodies; MSA, multiple system atrophy; PSP, progressive supranuclear palsy; CBD, corticobasal degeneration.

Supplementary Table 3: Individual feature-diagnosis respondent attribution

| Clinical Feature | Diagnosis | Framework association strength | Respondents,  n (%) [95% CI] |
| --- | --- | --- | --- |
| Orthostatic lightheadedness or syncope | CBD | 0 | 0 (0.0%) [0.0, 11.0] |
| Orthostatic lightheadedness or syncope | DLB | 1+ | 15 (48.4%) [32.0, 65.2] |
| Orthostatic lightheadedness or syncope | MSA | 2+ | 31 (100.0%) [89.0, 100.0] |
| Orthostatic lightheadedness or syncope | PSP | 0 | 0 (0.0%) [0.0, 11.0] |
| Asymmetric loss of “coordination” or alien limb | CBD | 2+ | 31 (100.0%) [89.0, 100.0] |
| Asymmetric loss of “coordination” or alien limb | DLB | 0 | 0 (0.0%) [0.0, 11.0] |
| Asymmetric loss of “coordination” or alien limb | MSA | 0 | 1 (3.2%) [0.6, 16.2] |
| Asymmetric loss of “coordination” or alien limb | PSP | 0 | 1 (3.2%) [0.6, 16.2] |
| Ataxia otherwise unexplained | CBD | 0 | 2 (6.5%) [1.8, 20.7] |
| Ataxia otherwise unexplained | DLB | 0 | 0 (0.0%) [0.0, 11.0] |
| Ataxia otherwise unexplained | MSA | 2+ | 31 (100.0%) [89.0, 100.0] |
| Ataxia otherwise unexplained | PSP | 0 | 2 (6.5%) [1.8, 20.7] |
| Dysphagia within 3 y of motor onset | CBD | 0 | 14 (45.2%) [29.2, 62.2] |
| Dysphagia within 3 y of motor onset | DLB | 0 | 2 (6.5%) [1.8, 20.7] |
| Dysphagia within 3 y of motor onset | MSA | 1+ | 25 (80.6%) [63.7, 90.8] |
| Dysphagia within 3 y of motor onset | PSP | 2+ | 24 (77.4%) [60.2, 88.6] |
| Unexplained falls within 3 y of onset | CBD | 0 | 11 (35.5%) [21.1, 53.1] |
| Unexplained falls within 3 y of onset | DLB | 0 | 9 (29.0%) [16.1, 46.6] |
| Unexplained falls within 3 y of onset | MSA | 1+ | 19 (61.3%) [43.8, 76.3] |
| Unexplained falls within 3 y of onset | PSP | 2+ | 31 (100.0%) [89.0, 100.0] |
| Nonrefractible blurring or diplopia | CBD | 0 | 4 (12.9%) [5.1, 28.9] |
| Nonrefractible blurring or diplopia | DLB | 0 | 1 (3.2%) [0.6, 16.2] |
| Nonrefractible blurring or diplopia | MSA | 1+ | 6 (19.4%) [9.2, 36.3] |
| Nonrefractible blurring or diplopia | PSP | 2+ | 29 (93.5%) [79.3, 98.2] |
| Stridor | CBD | 0 | 0 (0.0%) [0.0, 11.0] |
| Stridor | DLB | 0 | 0 (0.0%) [0.0, 11.0] |
| Stridor | MSA | 2+ | 29 (93.5%) [79.3, 98.2] |
| Stridor | PSP | 0 | 2 (6.5%) [1.8, 20.7] |
| Unexplained autonomic issues besides neurogenic orthostatic hypotension (nOH), such as urinary changes, constipation, ED, temperature intolerance | CBD | 0 | 3 (9.7%) [3.4, 24.9] |
| Unexplained autonomic issues besides neurogenic orthostatic hypotension (nOH), such as urinary changes, constipation, ED, temperature intolerance | DLB | 1+ | 13 (41.9%) [26.4, 59.2] |
| Unexplained autonomic issues besides neurogenic orthostatic hypotension (nOH), such as urinary changes, constipation, ED, temperature intolerance | MSA | 2+ | 30 (96.8%) [83.8, 99.4] |
| Unexplained autonomic issues besides neurogenic orthostatic hypotension (nOH), such as urinary changes, constipation, ED, temperature intolerance | PSP | 0 | 4 (12.9%) [5.1, 28.9] |
| Dream enactment without recall (RBD) | CBD | 0 | 0 (0.0%) [0.0, 11.0] |
| Dream enactment without recall (RBD) | DLB | 2+ | 31 (100.0%) [89.0, 100.0] |
| Dream enactment without recall (RBD) | MSA | 2+ | 26 (83.9%) [67.4, 92.9] |
| Dream enactment without recall (RBD) | PSP | 0 | 2 (6.5%) [1.8, 20.7] |
| Speech and language impairment | CBD | 2+ | 27 (87.1%) [71.2, 94.9] |
| Speech and language impairment | DLB | 2+ | 18 (58.1%) [40.8, 73.6] |
| Speech and language impairment | MSA | 0 | 16 (51.6%) [34.8, 68.0] |
| Speech and language impairment | PSP | 1+ | 24 (77.4%) [60.2, 88.6] |
| Visual hallucinations not otherwise explained | CBD | 0 | 2 (6.5%) [1.8, 20.7] |
| Visual hallucinations not otherwise explained | DLB | 2+ | 31 (100.0%) [89.0, 100.0] |
| Visual hallucinations not otherwise explained | MSA | 0 | 1 (3.2%) [0.6, 16.2] |
| Visual hallucinations not otherwise explained | PSP | 0 | 1 (3.2%) [0.6, 16.2] |
| Major vascular risk factors or stroke history | CBD | 0 | 4 (12.9%) [5.1, 28.9] |
| Major vascular risk factors or stroke history | DLB | 0 | 1 (3.2%) [0.6, 16.2] |
| Major vascular risk factors or stroke history | MSA | 0 | 0 (0.0%) [0.0, 11.0] |
| Major vascular risk factors or stroke history | PSP | 0 | 2 (6.5%) [1.8, 20.7] |
| Modest, short-lived or no levodopa benefit | CBD | 0 | 28 (90.3%) [75.1, 96.7] |
| Modest, short-lived or no levodopa benefit | DLB | 2+ | 13 (41.9%) [26.4, 59.2] |
| Modest, short-lived or no levodopa benefit | MSA | 1+ | 28 (90.3%) [75.1, 96.7] |
| Modest, short-lived or no levodopa benefit | PSP | 1+ | 29 (93.5%) [79.3, 98.2] |
| Recent or current neuroleptic or other dopamine-blocking drug | CBD | 0 | 1 (3.2%) [0.6, 16.2] |
| Recent or current neuroleptic or other dopamine-blocking drug | DLB | 0 | 9 (29.0%) [16.1, 46.6] |
| Recent or current neuroleptic or other dopamine-blocking drug | MSA | 0 | 0 (0.0%) [0.0, 11.0] |
| Recent or current neuroleptic or other dopamine-blocking drug | PSP | 0 | 2 (6.5%) [1.8, 20.7] |
| Neurogenic orthostatic hypotension (nOH), especially without compensatory tachycardia | CBD | 0 | 0 (0.0%) [0.0, 11.0] |
| Neurogenic orthostatic hypotension (nOH), especially without compensatory tachycardia | DLB | 2+ | 13 (41.9%) [26.4, 59.2] |
| Neurogenic orthostatic hypotension (nOH), especially without compensatory tachycardia | MSA | 2+ | 31 (100.0%) [89.0, 100.0] |
| Neurogenic orthostatic hypotension (nOH), especially without compensatory tachycardia | PSP | 0 | 0 (0.0%) [0.0, 11.0] |
| Balance loss disproportionate to Parkinsonism | CBD | 0 | 11 (35.5%) [21.1, 53.1] |
| Balance loss disproportionate to Parkinsonism | DLB | 1+ | 5 (16.1%) [7.1, 32.6] |
| Balance loss disproportionate to Parkinsonism | MSA | 2+ | 22 (71.0%) [53.4, 83.9] |
| Balance loss disproportionate to Parkinsonism | PSP | 2+ | 26 (83.9%) [67.4, 92.9] |
| Gait apraxia/frontal gait syndrome | CBD | 1+ | 17 (54.8%) [37.8, 70.8] |
| Gait apraxia/frontal gait syndrome | DLB | 0 | 0 (0.0%) [0.0, 11.0] |
| Gait apraxia/frontal gait syndrome | MSA | 0 | 2 (6.5%) [1.8, 20.7] |
| Gait apraxia/frontal gait syndrome | PSP | 1+ | 25 (80.6%) [63.7, 90.8] |
| Myoclonus | CBD | 2+ | 24 (77.4%) [60.2, 88.6] |
| Myoclonus | DLB | 1+ | 9 (29.0%) [16.1, 46.6] |
| Myoclonus | MSA | 1+ | 13 (41.9%) [26.4, 59.2] |
| Myoclonus | PSP | 0 | 4 (12.9%) [5.1, 28.9] |
| Wide-based or ataxic gait without other obvious explanation | CBD | 0 | 1 (3.2%) [0.6, 16.2] |
| Wide-based or ataxic gait without other obvious explanation | DLB | 0 | 0 (0.0%) [0.0, 11.0] |
| Wide-based or ataxic gait without other obvious explanation | MSA | 2+ | 29 (93.5%) [79.3, 98.2] |
| Wide-based or ataxic gait without other obvious explanation | PSP | 1+ | 3 (9.7%) [3.4, 24.9] |
| Apraxia of up or down saccades (difficulty initiating movement) | CBD | 2+ | 11 (35.5%) [21.1, 53.1] |
| Apraxia of up or down saccades (difficulty initiating movement) | DLB | 0 | 0 (0.0%) [0.0, 11.0] |
| Apraxia of up or down saccades (difficulty initiating movement) | MSA | 0 | 0 (0.0%) [0.0, 11.0] |
| Apraxia of up or down saccades (difficulty initiating movement) | PSP | 1+ | 30 (96.8%) [83.8, 99.4] |
| Blepharospasm, excessive blinking, or photophobia | CBD | 0 | 2 (6.5%) [1.8, 20.7] |
| Blepharospasm, excessive blinking, or photophobia | DLB | 0 | 0 (0.0%) [0.0, 11.0] |
| Blepharospasm, excessive blinking, or photophobia | MSA | 1+ | 3 (9.7%) [3.4, 24.9] |
| Blepharospasm, excessive blinking, or photophobia | PSP | 2+ | 26 (83.9%) [67.4, 92.9] |
| Equally reduced upward and downward gaze | CBD | 1+ | 3 (9.7%) [3.4, 24.9] |
| Equally reduced upward and downward gaze | DLB | 2+ | 0 (0.0%) [0.0, 11.0] |
| Equally reduced upward and downward gaze | MSA | 1+ | 0 (0.0%) [0.0, 11.0] |
| Equally reduced upward and downward gaze | PSP | 0 | 27 (87.1%) [71.2, 94.9] |
| Square wave jerks, large and/or frequent | CBD | 0 | 1 (3.2%) [0.6, 16.2] |
| Square wave jerks, large and/or frequent | DLB | 0 | 0 (0.0%) [0.0, 11.0] |
| Square wave jerks, large and/or frequent | MSA | 1+ | 11 (35.5%) [21.1, 53.1] |
| Square wave jerks, large and/or frequent | PSP | 2+ | 25 (80.6%) [63.7, 90.8] |
| Vertical > horizontal gaze palsy, reduced speed/size of downward saccades, or curved downward saccades | CBD | 1+ | 1 (3.2%) [0.6, 16.2] |
| Vertical > horizontal gaze palsy, reduced speed/size of downward saccades, or curved downward saccades | DLB | 0 | 0 (0.0%) [0.0, 11.0] |
| Vertical > horizontal gaze palsy, reduced speed/size of downward saccades, or curved downward saccades | MSA | 0 | 0 (0.0%) [0.0, 11.0] |
| Vertical > horizontal gaze palsy, reduced speed/size of downward saccades, or curved downward saccades | PSP | 2+ | 31 (100.0%) [89.0, 100.0] |
| Aphasia or other cognitive defects | CBD | 2+ | 28 (90.3%) [75.1, 96.7] |
| Aphasia or other cognitive defects | DLB | 0 | 15 (48.4%) [32.0, 65.2] |
| Aphasia or other cognitive defects | MSA | 0 | 3 (9.7%) [3.4, 24.9] |
| Aphasia or other cognitive defects | PSP | 0 | 19 (61.3%) [43.8, 76.3] |
| Impulsivity, manipulating objects | CBD | 0 | 18 (58.1%) [40.8, 73.6] |
| Impulsivity, manipulating objects | DLB | 0 | 6 (19.4%) [9.2, 36.3] |
| Impulsivity, manipulating objects | MSA | 1+ | 0 (0.0%) [0.0, 11.0] |
| Impulsivity, manipulating objects | PSP | 2+ | 23 (74.2%) [56.8, 86.3] |
| Palilalia | CBD | 0 | 15 (48.4%) [32.0, 65.2] |
| Palilalia | DLB | 0 | 2 (6.5%) [1.8, 20.7] |
| Palilalia | MSA | 0 | 2 (6.5%) [1.8, 20.7] |
| Palilalia | PSP | 2+ | 23 (74.2%) [56.8, 86.3] |
| Anterocollis disproportionate to Parkinsonism | CBD | 0 | 0 (0.0%) [0.0, 11.0] |
| Anterocollis disproportionate to Parkinsonism | DLB | 1+ | 0 (0.0%) [0.0, 11.0] |
| Anterocollis disproportionate to Parkinsonism | MSA | 2+ | 30 (96.8%) [83.8, 99.4] |
| Anterocollis disproportionate to Parkinsonism | PSP | 0 | 3 (9.7%) [3.4, 24.9] |
| Asymmetric astereognosis | CBD | 2+ | 31 (100.0%) [89.0, 100.0] |
| Asymmetric astereognosis | DLB | 1+ | 1 (3.2%) [0.6, 16.2] |
| Asymmetric astereognosis | MSA | 0 | 1 (3.2%) [0.6, 16.2] |
| Asymmetric astereognosis | PSP | 0 | 3 (9.7%) [3.4, 24.9] |
| Asymmetric dystonia | CBD | 2+ | 29 (93.5%) [79.3, 98.2] |
| Asymmetric dystonia | DLB | 0 | 1 (3.2%) [0.6, 16.2] |
| Asymmetric dystonia | MSA | 1+ | 5 (16.1%) [7.1, 32.6] |
| Asymmetric dystonia | PSP | 1+ | 4 (12.9%) [5.1, 28.9] |
| Asymmetric limb apraxia | CBD | 2+ | 31 (100.0%) [89.0, 100.0] |
| Asymmetric limb apraxia | DLB | 0 | 0 (0.0%) [0.0, 11.0] |
| Asymmetric limb apraxia | MSA | 0 | 1 (3.2%) [0.6, 16.2] |
| Asymmetric limb apraxia | PSP | 1+ | 3 (9.7%) [3.4, 24.9] |
| Asymmetric pyramidal signs | CBD | 2+ | 30 (96.8%) [83.8, 99.4] |
| Asymmetric pyramidal signs | DLB | 0 | 0 (0.0%) [0.0, 11.0] |
| Asymmetric pyramidal signs | MSA | 0 | 9 (29.0%) [16.1, 46.6] |
| Asymmetric pyramidal signs | PSP | 0 | 5 (16.1%) [7.1, 32.6] |

Caption: For each of the 128 clinical feature-diagnosis combinations, respondent attribution is shown alongside the corresponding association strength in the published framework. Values represent the number and percentage of 31 movement disorders specialist respondents who selected the diagnosis, with 95% Wilson score confidence intervals. DLB, dementia with Lewy bodies; MSA, multiple system atrophy; PSP, progressive supranuclear palsy; CBD, corticobasal degeneration.

Supplementary Table 4: Feature-level exact-set agreement using moderate (2+) associations

| Clinical Feature | Framework Mapped Diagnoses (2+) | Exact-set agreement,  n (%) [95% CI] † |
| --- | --- | --- |
| Vertical > horizontal gaze palsy, reduced speed/size of downward saccades, or curved downward saccades (E) | PSP | 30 (96.8%) [83.8, 99.4] |
| Asymmetric loss of “coordination” or alien limb (H) | CBD | 29 (93.5%) [79.3, 98.2] |
| Visual hallucinations not otherwise explained (H) | DLB | 29 (93.5%) [79.3, 98.2] |
| Anterocollis disproportionate to Parkinsonism (E) | MSA | 28 (90.3%) [75.1, 96.7] |
| Ataxia otherwise unexplained (H) | MSA | 28 (90.3%) [75.1, 96.7] |
| Asymmetric astereognosis (E) | CBD | 27 (87.1%) [71.1, 94.9] |
| Asymmetric limb apraxia (E) | CBD | 27 (87.1%) [71.1, 94.9] |
| Stridor (H) | MSA | 27 (87.1%) [71.1, 94.9] |
| Wide-based or ataxic gait without other obvious explanation (E) | MSA | 26 (83.9%) [67.4, 92.9] |
| Dream enactment without recall (RBD) (H) | DLB, MSA | 24 (77.4%) [60.2, 88.6] |
| Major vascular risk factors or stroke history (H) | None | 24 (77.4%) [60.2, 88.6] |
| Nonrefractible blurring or diplopia (H) | PSP | 22 (71.0%) [53.4, 83.9] |
| Blepharospasm, excessive blinking, or photophobia (E) | PSP | 22 (71.0%) [53.4, 83.9] |
| Asymmetric dystonia (E) | CBD | 22 (71.0%) [53.4, 83.9] |
| Asymmetric pyramidal signs (E) | CBD | 21 (67.7%) [50.1, 81.4] |
| Recent or current neuroleptic or other dopamine-blocking drug (H) | None | 20 (64.5%) [46.9, 78.9] |
| Square wave jerks, large and/or frequent (E) | PSP | 19 (61.3%) [43.8, 76.3] |
| Unexplained autonomic issues besides neurogenic orthostatic hypotension (nOH), such as urinary changes, constipation, ED, temperature intolerance (H) | MSA | 17 (54.8%) [37.8, 70.8] |
| Orthostatic lightheadedness or syncope (H) | MSA | 16 (51.6%) [34.8, 68.0] |
| Neurogenic orthostatic hypotension (nOH) especially without compensatory tachycardia (E) | DLB, MSA | 13 (41.9%) [26.4, 59.2] |
| Myoclonus (E) | CBD | 10 (32.3%) [18.6, 49.9] |
| Palilalia (E) | PSP | 10 (32.3%) [18.6, 49.9] |
| Impulsivity, manipulating objects (E) | PSP | 9 (29.0%) [16.1, 46.6] |
| Unexplained falls within 3 y of onset (H) | PSP | 8 (25.8%) [13.7, 43.2] |
| Aphasia or other cognitive defects (E) | CBD | 7 (22.6%) [11.4, 39.8] |
| Balance loss disproportionate to Parkinsonism (E) | MSA, PSP | 7 (22.6%) [11.4, 39.8] |
| Gait apraxia/frontal gait syndrome (E) | None | 3 (9.7%) [3.3, 24.9] |
| Dysphagia within 3 y of motor onset (H) | PSP | 2 (6.5%) [1.8, 20.7] |
| Apraxia of up or down saccades (difficulty initiating movement) (E) | CBD | 1 (3.2%) [0.6, 16.2] |
| Speech and language impairment (H) | CBD, DLB | 1 (3.2%) [0.6, 16.2] |
| Modest, short-lived or no levodopa benefit (H) | DLB | 0 (0.0%) [0.0, 11.0] |
| Equally reduced upward and downward gaze (E) | DLB | 0 (0.0%) [0.0, 11.0] |

Caption: Feature-level exact-set agreement with the moderate (2+) threshold mapping among 31 movement disorders specialist respondents. Exact-set agreement required the respondent to select all and only diagnoses assigned a moderate (2+) association with the clinical feature in the published framework. Omission of a mapped diagnosis or selection of an additional diagnosis was classified as non-agreement. Features without a moderate association with any of the four APDs were represented by an empty reference set (None); respondents matched that reference set only when no APD was selected. Values are reported as n (%) with 95% Wilson score confidence intervals. DLB, dementia with Lewy bodies; MSA, multiple system atrophy; PSP, progressive supranuclear palsy; CBD, corticobasal degeneration.

Supplementary Table 5: Feature-level exact-set agreement using all mapped (1+ and 2+) associations

| Clinical Feature | Framework Mapped Diagnoses (1+/2+) | Exact-set agreement,  n (%) [95% CI] |
| --- | --- | --- |
| Asymmetric loss of “coordination” or alien limb (H) | CBD | 29 (93.5%) [79.3, 98.2] |
| Visual hallucinations not otherwise explained (H) | DLB | 29 (93.5%) [79.3, 98.2] |
| Ataxia otherwise unexplained (H) | MSA | 28 (90.3%) [75.1, 96.7] |
| Stridor (H) | MSA | 27 (87.1%) [71.1, 94.9] |
| Dream enactment without recall (RBD) (H) | DLB, MSA | 24 (77.4%) [60.2, 88.6] |
| Major vascular risk factors or stroke history (H) | None | 24 (77.4%) [60.2, 88.6] |
| Asymmetric pyramidal signs (E) | CBD | 21 (67.7%) [50.1, 81.4] |
| Recent or current neuroleptic or other dopamine-blocking drug (H) | None | 20 (64.5%) [46.9, 78.9] |
| Orthostatic lightheadedness or syncope (H) | DLB, MSA | 15 (48.4%) [32.0, 65.2] |
| Gait apraxia/frontal gait syndrome (E) | PSP, CBD | 14 (45.2%) [29.2, 62.2] |
| Neurogenic orthostatic hypotension (nOH) especially without compensatory tachycardia (E) | DLB, MSA | 13 (41.9%) [26.4, 59.2] |
| Dysphagia within 3 y of motor onset (H) | MSA, PSP | 10 (32.3%) [18.6, 49.9] |
| Apraxia of up or down saccades (difficulty initiating movement) (E) | PSP, CBD | 10 (32.3%) [18.6, 49.9] |
| Palilalia (E) | PSP | 10 (32.3%) [18.6, 49.9] |
| Unexplained falls within 3 y of onset (H) | MSA, PSP | 9 (29.0%) [16.1, 46.6] |
| Unexplained autonomic issues besides neurogenic orthostatic hypotension (nOH), such as urinary changes, constipation, ED, temperature intolerance (H) | DLB, MSA | 9 (29.0%) [16.1, 46.6] |
| Aphasia or other cognitive defects (E) | CBD | 7 (22.6%) [11.4, 39.8] |
| Square wave jerks, large and/or frequent (E) | MSA, PSP | 5 (16.1%) [7.1, 32.6] |
| Speech and language impairment (H) | DLB, PSP, CBD | 3 (9.7%) [3.3, 24.9] |
| Nonrefractible blurring or diplopia (H) | MSA, PSP | 3 (9.7%) [3.3, 24.9] |
| Asymmetric limb apraxia (E) | PSP, CBD | 3 (9.7%) [3.3, 24.9] |
| Wide-based or ataxic gait without other obvious explanation (E) | MSA, PSP | 2 (6.5%) [1.8, 20.7] |
| Blepharospasm, excessive blinking, or photophobia (E) | MSA, PSP | 2 (6.5%) [1.8, 20.7] |
| Myoclonus (E) | DLB, MSA, CBD | 2 (6.5%) [1.8, 20.7] |
| Asymmetric astereognosis (E) | DLB, CBD | 1 (3.2%) [0.6, 16.2] |
| Vertical > horizontal gaze palsy, reduced speed/size of downward saccades, or curved downward saccades (E) | PSP, CBD | 1 (3.2%) [0.6, 16.2] |
| Asymmetric dystonia (E) | MSA, PSP, CBD | 1 (3.2%) [0.6, 16.2] |
| Modest, short-lived or no levodopa benefit (H) | DLB, MSA, PSP | 0 (0.0%) [0.0, 11.0] |
| Balance loss disproportionate to Parkinsonism (E) | DLB, MSA, PSP | 0 (0.0%) [0.0, 11.0] |
| Equally reduced upward and downward gaze (E) | DLB, MSA, CBD | 0 (0.0%) [0.0, 11.0] |
| Anterocollis disproportionate to Parkinsonism (E) | DLB, MSA | 0 (0.0%) [0.0, 11.0] |
| Impulsivity, manipulating objects (E) | MSA, PSP | 0 (0.0%) [0.0, 11.0] |

Caption: Feature-level exact-set agreement with the all-mapped (1+/2+) threshold mapping among 31 movement disorders specialist respondents. Exact-set agreement required the respondent to select all and only diagnoses assigned either a modest (1+) or moderate (2+) association with the clinical feature in the published framework. Omission of a mapped diagnosis or selection of an additional diagnosis was classified as non-agreement. Features without a modest (1+) or moderate (2+) association with any of the four APDs were represented by an empty reference set (None); respondents matched that reference set only when no APD was selected. Values are reported as n (%) with 95% Wilson score confidence intervals. DLB, dementia with Lewy bodies; MSA, multiple system atrophy; PSP, progressive supranuclear palsy; CBD, corticobasal degeneration.

Supplementary Table 6: Exact-set agreement sensitivity analysis excluding the corticobasal diagnostic category

| Clinical Feature | Exact-set agreement, n (%) [95% CI] | |
| --- | --- | --- |
|  | **2+ mapping** | **1+/2+ mapping** |
| Vertical > horizontal gaze palsy, reduced speed/size of downward saccades, or curved downward saccades | 31 (100.0%) [89.0, 100.0] | 31 (100.0%) [89.0, 100.0] |
| Visual hallucinations not otherwise explained | 30 (96.8%) [83.8, 99.4] | 30 (96.8%) [83.8, 99.4] |
| Ataxia otherwise unexplained | 29 (93.5%) [79.3, 98.2] | 29 (93.5%) [79.3, 98.2] |
| Asymmetric loss of “coordination” or alien limb | 29 (93.5%) [79.3, 98.2] | 29 (93.5%) [79.3, 98.2] |
| Anterocollis disproportionate to Parkinsonism | 28 (90.3%) [75.1, 96.7] | 0 (0.0%) [0.0, 11.0] |
| Major vascular risk factors or stroke history | 28 (90.3%) [75.1, 96.7] | 28 (90.3%) [75.1, 96.7] |
| Stridor | 27 (87.1%) [71.1, 94.9] | 27 (87.1%) [71.1, 94.9] |
| Asymmetric astereognosis | 27 (87.1%) [71.1, 94.9] | 1 (3.2%) [0.6, 16.2] |
| Wide-based or ataxic gait without other obvious explanation | 27 (87.1%) [71.1, 94.9] | 2 (6.5%) [1.8, 20.7] |
| Asymmetric limb apraxia | 27 (87.1%) [71.1, 94.9] | 3 (9.7%) [3.3, 24.9] |
| Nonrefractible blurring or diplopia | 25 (80.6%) [63.7, 90.8] | 3 (9.7%) [3.3, 24.9] |
| Blepharospasm, excessive blinking, or photophobia | 24 (77.4%) [60.2, 88.6] | 2 (6.5%) [1.8, 20.7] |
| Dream enactment without recall (RBD) | 24 (77.4%) [60.2, 88.6] | 24 (77.4%) [60.2, 88.6] |
| Asymmetric dystonia | 22 (71.0%) [53.4, 83.9] | 1 (3.2%) [0.6, 16.2] |
| Asymmetric pyramidal signs | 22 (71.0%) [53.4, 83.9] | 22 (71.0%) [53.4, 83.9] |
| Recent or current neuroleptic or other dopamine-blocking drug | 20 (64.5%) [46.9, 78.9] | 20 (64.5%) [46.9, 78.9] |
| Square wave jerks, large and/or frequent | 20 (64.5%) [46.9, 78.9] | 5 (16.1%) [7.1, 32.6] |
| Palilalia | 19 (61.3%) [43.8, 76.3] | 19 (61.3%) [43.8, 76.3] |
| Impulsivity, manipulating objects | 18 (58.1%) [40.8, 73.6] | 0 (0.0%) [0.0, 11.0] |
| Unexplained autonomic issues besides neurogenic orthostatic hypotension (nOH), such as urinary changes, constipation, ED, temperature intolerance | 17 (54.8%) [37.8, 70.8] | 9 (29.0%) [16.1, 46.6] |
| Orthostatic lightheadedness or syncope | 16 (51.6%) [34.8, 68.0] | 15 (48.4%) [32.0, 65.2] |
| Neurogenic orthostatic hypotension (nOH), especially without compensatory tachycardia | 13 (41.9%) [26.4, 59.2] | 13 (41.9%) [26.4, 59.2] |
| Balance loss disproportionate to Parkinsonism | 12 (38.7%) [23.7, 56.2] | 5 (16.1%) [7.1, 32.6] |
| Myoclonus | 12 (38.7%) [23.7, 56.2] | 3 (9.7%) [3.3, 24.9] |
| Unexplained falls within 3 y of onset | 10 (32.3%) [18.6, 49.9] | 12 (38.7%) [23.7, 56.2] |
| Aphasia or other cognitive defects | 8 (25.8%) [13.7, 43.2] | 8 (25.8%) [13.7, 43.2] |
| Gait apraxia/frontal gait syndrome | 5 (16.1%) [7.1, 32.6] | 24 (77.4%) [60.2, 88.6] |
| Dysphagia within 3 y of motor onset (H) | 4 (12.9%) [5.1, 28.9] | 18 (58.1%) [40.8, 73.6] |
| Speech and language impairment | 3 (9.7%) [3.3, 24.9] | 3 (9.7%) [3.3, 24.9] |
| Apraxia of up or down saccades (difficulty initiating movement) | 1 (3.2%) [0.6, 16.2] | 30 (96.8%) [83.8, 99.4] |
| Modest, short-lived or no levodopa benefit | 0 (0.0%) [0.0, 11.0] | 13 (41.9%) [26.4, 59.2] |
| Equally reduced upward and downward gaze | 0 (0.0%) [0.0, 11.0] | 0 (0.0%) [0.0, 11.0] |

Caption: Sensitivity analysis of feature-level exact-set agreement among 31 movement disorders specialist respondents after excluding the corticobasal diagnostic category from both respondent selections and framework-derived reference sets. Exact-set agreement is reported under the moderate-association (2+-only) mapping and the all-mapped (1+/2+) mapping. Values are n (%) with 95% Wilson score confidence intervals.

Supplementary Table 7: Non-movement disorders inter-rater agreement in feature-diagnosis attribution, overall and by diagnosis

| Analysis set | Feature-diagnosis pairs, n | Raw agreement (%) | Gwet's AC1 [95% CI] |
| --- | --- | --- | --- |
| Overall | 128 | 71.5 | 0.53 [0.49, 0.56] |
| DLB | 32 | 67.3 | 0.45[0.40, 0.50] |
| MSA | 32 | 75.7 | 0.62 [0.54, 0.70] |
| PSP | 32 | 70.4 | 0.47 [0.41, 0.53] |
| CBD | 32 | 72.5 | 0.55 [0.48, 0.62] |

Caption: Inter-specialist agreement in binary feature-diagnosis attribution among 14 non-movement disorders specialist respondents including: Behavioral neurologists (n = 4), Other neurologists (n = 8), and Nurse practitioners (n = 2). Each of the 128 feature-diagnosis pairs (32 clinical features × four APD diagnoses) was treated as an item rated independently by all respondents. Raw percent agreement and Gwet's AC1 with 95% confidence intervals are reported for the complete set of feature-diagnosis pairs and separately for each diagnosis. DLB, dementia with Lewy bodies; MSA, multiple system atrophy; PSP, progressive supranuclear palsy; CBD, corticobasal degeneration.

Supplementary Table 8: Diagnosis-level coverage of framework-mapped clinical features by non-movement disorders specialist respondents

| Diagnosis | Moderate (2+) associations, n | Coverage,  median (IQR) | All mapped (1+/2+) associations, n | Coverage,  median (IQR) |
| --- | --- | --- | --- | --- |
| DLB | 6 | 58.3% (50.0–79.2) | 12 | 45.8% (27.1–50.0) |
| MSA | 9 | 55.6% (47.2–66.7) | 19 | 36.8% (31.6–42.1) |
| PSP | 9 | 66.7% (38.9–86.1) | 16 | 56.2% (40.6–71.9) |
| CBD | 9 | 55.6% (38.9–83.3) | 12 | 41.7% (29.2–64.6) |

Caption: Coverage represents, for each non-movement disorders specialist respondent and diagnosis, the proportion of clinical features mapped to that diagnosis in the published framework that were selected by the respondent. Additional diagnoses selected for an individual feature did not reduce coverage. Moderate-association coverage includes only framework associations designated 2+; All mapped association coverage includes associations designated either 1+ or 2+. Values represent the median and interquartile range across 14 non-movement disorders specialist respondents. DLB, dementia with Lewy bodies; MSA, multiple system atrophy; PSP, progressive supranuclear palsy; CBD, corticobasal degeneration.

**SUPPLEMENTARY FIGURES**

Supplementary Figure 1: Familiarity with APD diagnostic criteria and the General Neurologist’s Algorithm
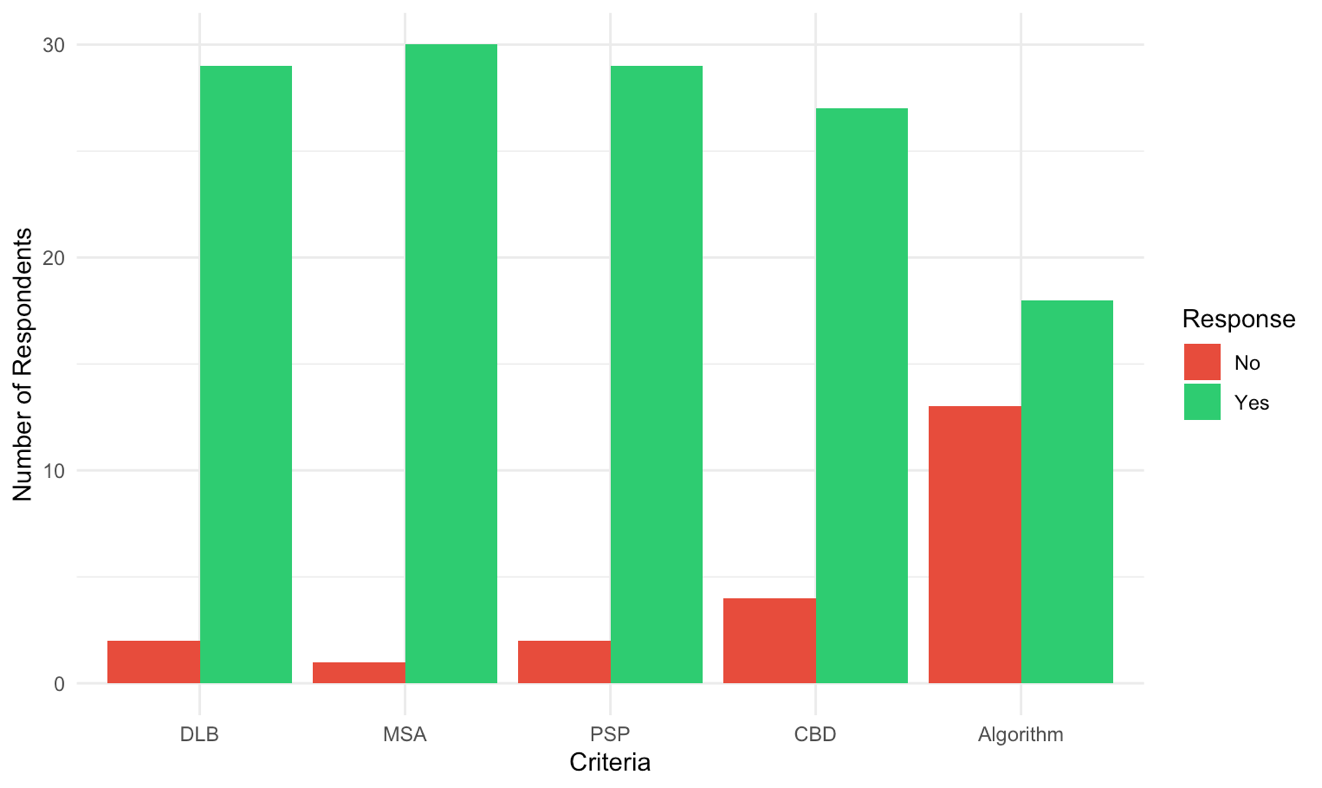

Caption: Movement disorders specialist respondents' reported familiarity with the four individual APD diagnostic criteria and the General Neurologist’s Algorithm (here, “Algorithm”). DLB, dementia with Lewy bodies; MSA, multiple system atrophy; PSP, progressive supranuclear palsy; CBD, corticobasal degeneration.

Supplementary Figure 2: Self-reported utilization of APD diagnostic criteria and the General Neurologist’s Algorithm
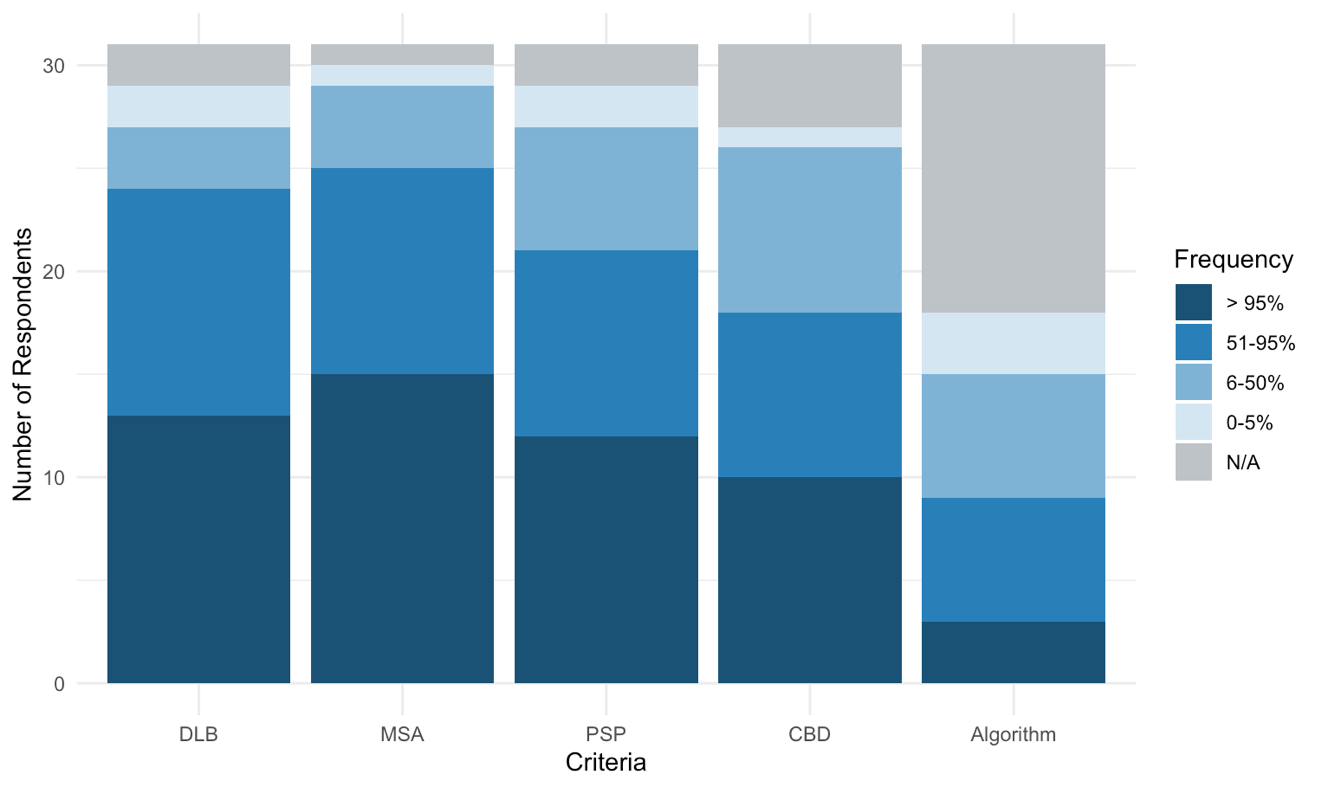

Caption: Movement disorders specialist respondents' reported frequency of use of the four individual APD diagnostic criteria and the General Neurologist’s Algorithm (here, “Algorithm”) in cases where the relevant APD is suspected. “N/A” refers to respondents who reported no familiarity with the corresponding criteria or algorithm and therefore did not provide usage data. DLB, dementia with Lewy bodies; MSA, multiple system atrophy; PSP, progressive supranuclear palsy; CBD, corticobasal degeneration.

Supplementary Figure 3: Self-reported likelihood of using an EHR-based application to apply APD criteria

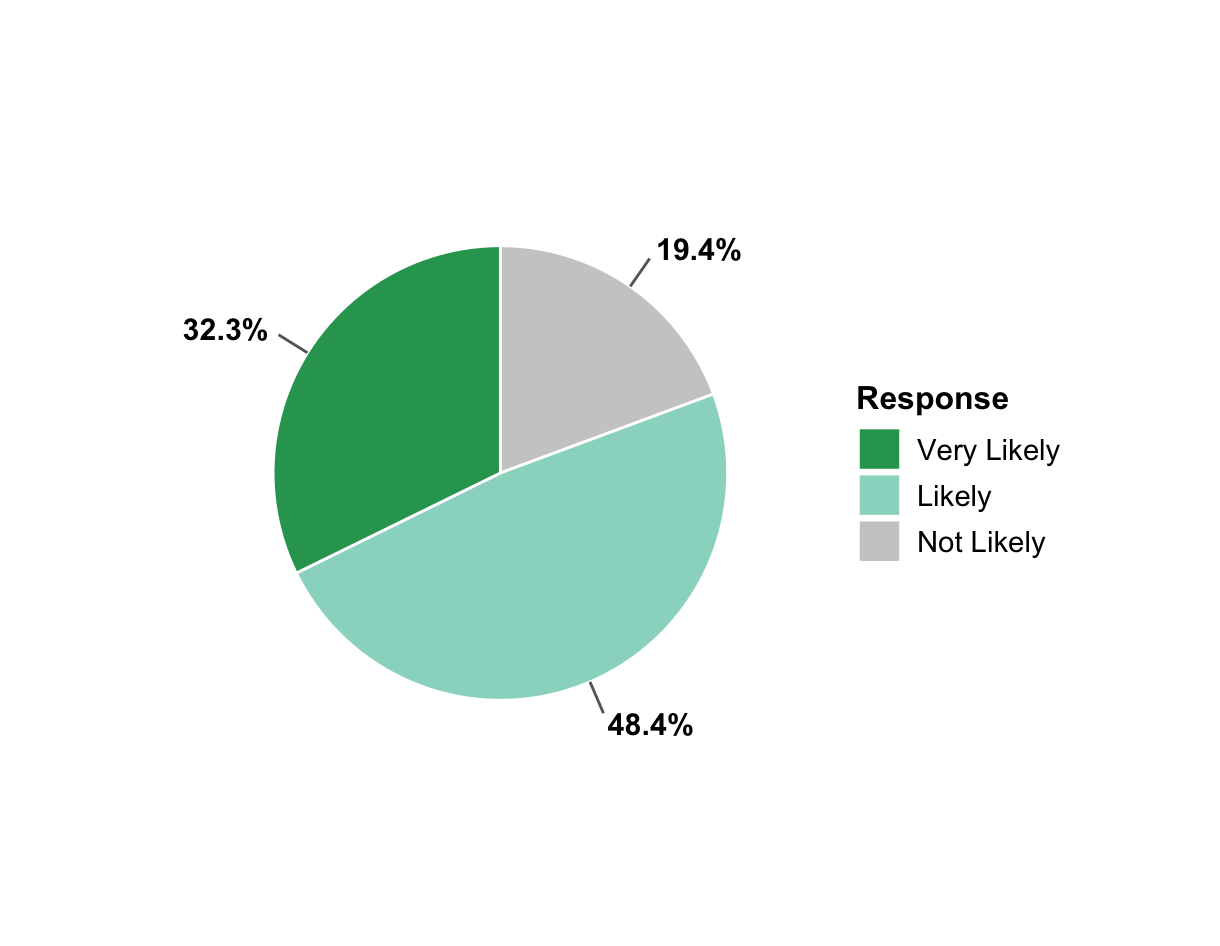

Caption: Movement disorders specialist respondents' self-reported likelihood of utilizing an EHR-based digital application to apply APD diagnostic criteria if it were made available.

Supplementary Figure 4: Perceived need for a unified diagnostic schema for APD

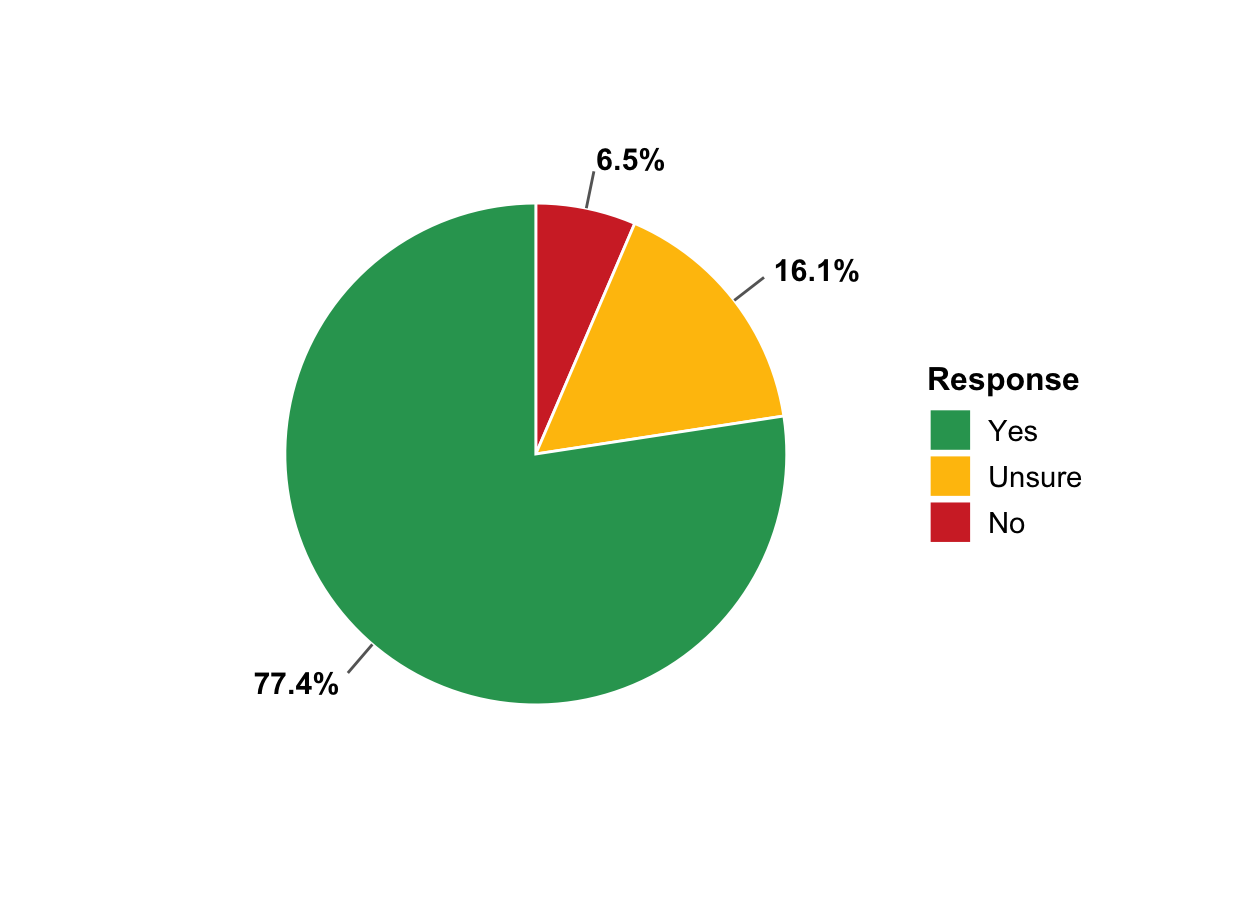

Caption: Movement disorders specialist respondents' opinions on whether a unified diagnostic schema that allows for application of multiple APD criteria in parallel is needed.
